# Impact of SARS-CoV-2 exposure history on antibody kinetics and correlates of protection in The Gambia

**DOI:** 10.64898/2026.01.02.26343369

**Authors:** David Hodgson, Rhys D. Wenlock, Natalie Barratt, Maries Gomes, Ya Jankey Jagne, Dawda Jobe, Sheikh Jarju, Madikoi Danso, Nigel Temperton, Beate Kampmann, Stefan Flasche, Adam Kucharski, Thushan I. de Silva

## Abstract

**BACKGROUND:** Understanding SARS-CoV-2 antibody dynamics is critical for pandemic preparedness, particularly where population immunity has developed through high infection rates with minimal vaccination. Whether predominantly asymptomatic infections confer protective immunity and which biomarkers best predict protection in resource-limited settings remain unclear.

**METHODS:** We conducted a household cohort study in The Gambia over 15 months (March 2021–June 2022) during Delta and Omicron waves, with weekly upper respiratory tract sampling for SARS-CoV-2 PCR testing. Serum and mucosal samples were collected at baseline, 6, and 12 months. We measured serum neutralising antibodies and mucosal IgA against five SARS-CoV-2 variants. Using Bayesian hierarchical models, we quantified antibody kinetics and evaluated correlates of protection by predictive accuracy across biomarker-variant combinations.

**FINDINGS:** Among 289 participants with 768 serological and mucosal measurements, attack rates of 53% (Delta) and 79% (Omicron) were observed, with 84% of infections asymptomatic. Delta infection produced serum neutralising antibody responses to the Delta variant (27-fold rise at peak, 95% CrI 5·0–95·2) persisting 197 days >4-fold baseline, while Delta mucosal IgA responses were shorter (9-fold rise at peak, 95% CrI 1·7–39·3, persisting 62 days). Delta infections generated substantial cross-reactive boosting to Ancestral and Alpha variants, whereas Omicron boosting was more specific to BA.1.

Participants with hybrid immunity from infection and vaccination achieved higher antibody levels (24-fold rise) than those with infection alone (6-fold rise) during the Delta wave. Variant-matched neutralisation demonstrated superior predicted protection performance against infection compared to other biomarkers in both waves (Delta: AUC 0·66; Omicron BA.1: AUC 0·65).

**INTERPRETATION:** Predominantly asymptomatic SARS-CoV-2 infections in The Gambia generated robust, durable and protective antibody responses. Variant-matched serum neutralising antibody levels capture protection as effectively as complex multi-biomarker approaches, providing pragmatic guidance for serological surveillance for population immunity in resource-constrained settings.

**FUNDING:** United Kingdom Research and Innovation Grant (No. MC_PC_19084).

**RESEARCH IN CONTEXT:** *Evidence before this study:* We searched PubMed Central for studies up to 29^th^ December 2025 using the terms (“SARS-CoV-2” OR “COVID-19”) AND “longitudinal serology” AND “sub-Saharan Africa”, with no language restrictions. This search yielded four results, of which only one was a longitudinal serological study conducted in sub-Saharan Africa that examined antibody dynamics and reinfection patterns in Kenya. The remaining three were a systematic review and meta-analysis on COVID-19 burden in developing countries, and two serological studies on non-COVID pathogens. In contrast, broader search term (“SARS-CoV-2” OR “COVID-19”) AND “longitudinal serology” returned 74 results, highlighting the stark underrepresentation of sub-Saharan African populations in longitudinal serology studies. This gap is critical because sub-Saharan African populations present distinct epidemiological features, including high reported rates of asymptomatic infections (65–85% compared to 20–35% globally), and population immunity predominantly driven by natural infection rather than vaccination. Further, most studies on correlates of protection have focused on vaccinated populations in high-income countries, using clinical trials or standard observational cohorts. Few have employed household-based design with intensive surveillance, which uniquely enables the identification of definite exposure events and precise temporal linkage between exposure and antibody measurements—overcoming a key limitation in which exposure timing is uncertain.

*Added value of this study:* This household-based cohort study provides high-resolution longitudinal immunological data from The Gambia across 15 months spanning Delta and Omicron waves, including 768 serological and mucosal measurements from 289 participants with intensive weekly PCR surveillance. We measured both serum neutralising antibodies and mucosal IgA against five SARS-CoV-2 variants concurrently, including both Ancestral and Alpha variants and contemporaneous circulating variants (Delta, Omicron BA.1, BA.2). The household design with defined exposure events and temporal antibody measurements provides a more consistent assessment of protection at the time of exposure compared to standard cohort studies. Using Bayesian hierarchical modelling, we quantified antibody kinetics following predominantly asymptomatic natural infections and identified optimal correlates of protection. We demonstrate that natural infection generates robust and durable antibody responses despite minimal symptoms, with Delta infection producing 18-fold rises in neutralising antibodies persisting 101 days above four-fold baseline. Importantly, we show that hybrid immunity, infection followed by vaccination, produces substantially higher antibody responses than infection alone, with 24-fold rises compared to 6-fold rises during the Delta wave. This hybrid immunity advantage was consistently maintained across subsequent exposures, demonstrating that vaccination provides meaningful immunological benefit even in highly infection-experienced populations. We show that prior infection history substantially influences antibody responses to antigenically similar variants through immune imprinting, whereas responses to the antigenically divergent Omicron variant were less affected by infection history. Critically, variant-matched serum pseudoneutralisation provided the correlate of protection with the best predictive capacity compared to other biomarkers and variant combinations, with Delta pseudoneutralisation optimal for the Delta wave (AUC 0·66) and Omicron BA.1 pseudoneutralisation optimal for the Omicron wave (AUC 0·65). Single functional serum neutralisation assays captured protection as effectively as more complex dual biomarker approaches combining serum and mucosal measurements.

*Implications of all the available evidence:* Our findings demonstrate that populations in sub-Saharan Africa with high natural infection rates and low vaccine coverage develop robust, durable immunity that provides baseline protection against future SARS-CoV-2 waves, even when infections are predominantly asymptomatic. Critically, however, the addition of vaccination to natural immunity produces substantially higher neutralising antibody levels, the optimal correlate of protection identified in this study, indicating that vaccination provides meaningful benefit even in highly seroprevalent populations with extensive prior infection exposure. The household-based design with intensive surveillance provides correlates of protection estimates that may be particularly relevant for assessing protection at defined exposure timepoints in community settings. The finding that single functional neutralisation assays, particularly those measuring variant-specific responses, perform comparably to complex multi-biomarker approaches provides pragmatic guidance for serological surveillance in resource-constrained settings, supporting investment in laboratory capacity for these assays while avoiding unnecessary complexity. The non-linear relationship between antibody levels and protection suggests that booster vaccination strategies can provide meaningful benefit even in highly seroprevalent populations. These insights from an underrepresented region inform evidence-based public health strategies and underscore the importance of conducting immunological research in diverse global contexts to achieve health equity and ensure pandemic preparedness.

## INTRODUCTION

The SARS-CoV-2 pandemic resulted in substantial morbidity and mortality, with over 704 million confirmed infections and seven million deaths reported worldwide as of January 2025.[1] Despite few reported COVID-19 cases in the first year, serological surveys across Africa revealed substantially higher SARS-CoV-2 seroprevalence, ranging from 22% to 29% in population-based studies.(1,2) By the end of the first Omicron wave, seroprevalence reached higher levels, with some studies suggesting near-universal seropositivity.(3) High infection rates with attenuated disease severity in sub-Saharan Africa, where asymptomatic infections comprise 65–85% of all SARS-CoV-2 infections,(4,5) compared to approximately 20–35% globally,(6) raised questions about factors influencing infection outcomes. Understanding antibody dynamics and correlates of protection in African contexts is essential for pandemic preparedness, yet data outside clinical trial settings remain limited.(7)

The emergence of antigenically divergent SARS-CoV-2 variants has shaped population immunity trajectories. In South Africa, the Omicron BA.1/BA.2 wave infected more than half the population, with reinfections and vaccine breakthroughs accounting for over 60% of infections, leaving less than 6% immunologically naïve.(8) Antigenic cartography studies demonstrate that while early variants cluster closely in antigenic space, Omicron linages are distinct antigenic outliers that escape immunity generated by prior variants,(9) raising questions about how infection history shapes antigenic responses to successive waves. In sub-Saharan Africa, high attack rates combined with larger proportions of asymptomatic infections, even after adjusting for age, have created distinct population immunity profiles.(3) Recent evidence suggests that vaccination can durably boost pre-existing infection-induced immunity in highly exposed populations,(10) yet the breadth and durability of antibody responses following natural infection alone, and the immunological consequences of repeated exposures in populations with minimal vaccine coverage, remain poorly characterised.(11,12) Understanding these immunological dynamics in African settings where natural infection is the primary driver of population immunity is critical for informing public health strategies.

To estimate the protective effects of the immunological response to SARS-CoV-2, the TRANSVIR study, a community-based cohort in The Gambia, longitudinally measured SARS-CoV-2 immune responses, and antibody kinetics across variant waves.(3,13) Previous findings from this cohort have highlighted intense transmission during the Delta and Omicron waves, occurring largely in the context of 14% population vaccination coverage in The Gambia. Furthermore, in March 2021, at the start of the study and prior to the emergence of the Delta variant, the household-adjusted SARS-CoV-2 seroprevalence in this Gambian population was 56.7% (95% CI: 50.0-63.0%). By the end of the study in June 2022, following the Delta and Omicron BA.1 waves, the household-adjusted seroprevalence had increased to 93.1% (95% CI: 86.0-96.7%).(13) Building on these insights, in this paper, we now focus on the dynamics of both systemic and mucosal antibodies across diverse exposure histories. We quantify how repeated exposures through infection shape the kinetics, durability, and variant-specific breadth of serum neutralising antibodies and mucosal IgA responses. By leveraging high-resolution longitudinal immunological data and Bayesian hierarchical modelling, we define how protective immune responses develop and change over time.

## METHODS

### PARTICIPANT RECRUITMENT, SAMPLING AND STUDY DESIGN

The TRANSVIR household study design has been previously described.(3,14) Following community and household sensitisation, families were invited to the MRCG clinical site for consenting. Recruited households ranged from 6–10 members, including at least one adult and one child. Written consent or thumbprint was obtained from all participants >18 years and assent was obtained from participants aged 12–18 years. Parents or guardians consented for participants under 12 years. Participants were followed up for 12 months with weekly combined throat and nose flocked swabs (TNS) collected into RNAprotect cell reagent (Qiagen).(3) Participants reported symptoms in the 7 days preceding each sampling timepoint. At enrollment and at 6-monthly visits, participants attended the clinic for venous blood sampling for serum separation and for collection of nasal lining fluid using a synthetic absorptive matrix (SAM) strip. If participants developed symptoms compatible with upper respiratory tract infection, they contacted the study team for an unscheduled visit. At unscheduled visits, the entire household was swabbed, with RT-PCR testing performed contemporaneously. Participants with a positive SARS-CoV-2 RT-PCR at unscheduled visits had an additional venous blood sample collected 28 days post-infection.

### ETHICS APPROVAL

Study approval was given by the joint Gambia Government and Medical Research Council Unit, The Gambia (MRCG) Ethics committee, and the London School of Hygiene and Tropical Medicine ethics committee (project ID 22556, clinicialtrials.gov NCT05952336).(14)Binding assays SARS-CoV-2 ELISA SARS-CoV-2 spike- (S) and N-specific immunoglobulin G (IgG) was measured using a previously described in-house ELISA, shown to have 99·5% sensitivity and 98.8% specificity for anti-S IgG, and 99·5% sensitivity and 84.1% specificity for anti-N IgG(15) Immunolon 4 HBX microplates (Thermo Scientic, USA) were coated with either the full-length extracellular domain of the SARS-CoV-2 spike glycoprotein produced in mammalian cells or a full-length N protein produced in Escherichia coli. A standard curve calibrated to the WHO International Standard for anti-SARS-CoV2 Immunoglobulin (NIBSC 20/136) was used to quantify S and N antibody titres. For each antigen, serostatus was determined using previously defined thresholds based on the ratio between the sample and control well optical densities. Pseudotype neutralisiation assay (pVNT)

Neutralisation assays were undertaken according to previously described methods.(16) Human serum was serially diluted (1:40 to 1:9720) in Ham’s F-12 and incubated with pseudoviruses representing ancestral, B.1.1.7 (Alpha), B.1.617.2 (Delta), and Omicron BA.1 and BA.2 variants (∼5ϑ×ϑ10^6^ RLU/well) for 1ϑh at 37ϑ°C. CHO-ACE2/TMPRSS2 cells (20,000/well) were then added and incubated for 48ϑh before luciferase assay. Internal calibrants and control wells (cell-only and serum-free) were included. Due to non-specific inhibition at serum dilutions below 1:40, samples with neutralisation titre (NT_50_) values below 1:40 were considered negative. The titre value is expressed as the serum dilution achieving 50% neutralisation (IC50).

#### Meso scale discovery assay to measure mucosal IgA

Mucosal anti-spike IgA responses to different SARS-CoV-2 spike variants (Ancestral, B.1.1.7, B.1.617.2, BA.1, BA.2) were measured using V-PLEX SARS-CoV-2 Panel 25 (IgA) Kit (K15585U, MSD, Maryland, USA) according to manufacturer’s instructions. To measure IgA binding antibodies, 96-well plates were blocked with Blocker A solution for 30ϑminutes. Plates were washed with MSD Wash buffer, and 50ϑμl of nasal mucosal lining fluid samples obtained from the SAM strips were diluted 1:25–1:800 and added to wells, along with standards and controls and incubated for 2ϑhours. After a 2-hour incubation and a wash step, diluted detection antibody (MSD SULFO-TAG™ Anti-Human IgA Antibody, was added and incubated for 1-hour. Plate was washed after the incubation and MSD GOLD Read Buffer B added. Plates were read using Discovery Bench 4.0 software provided by MESO SCALE DIAGNOSTICS, LLC. SAM strips processing for mucosal IgA measurements is previously described.(13) Readout units are expressed as AU/mL (arbitrary antibody units/millilitre) based on standard curves.

### MODELLING FRAMEWORK

We developed a Bayesian hierarchical model to jointly estimate antibody kinetics and correlates of protection for ten biomarkers (Ancestral, Alpha, Delta, Omicron BA.1, and Omicron BA.2 measured by pseudovirus neutralisation and mucosal anti-spike IgA binding) across both pandemic waves. This analysis was conducted as a post-hoc exploration of the data, extending beyond the pre-specified analysis plan outlined in the study protocol. The framework integrated: (1) antibody kinetics modelling waning and boosting dynamics following infection or vaccination, (2) a correlate of protection model relating log-antibody levels to infection risk given exposure, and (3) an exposure model accounting for household transmission intensity. (Full details in Methods S2–3)

Antibody trajectories were modelled on the log_10_ scale, with linear waning and parametric boosting post-exposure using power-function formulations adapted from Teunis et al.(17), comparing predicted to measured titres using Gaussian likelihoods with left-censoring at the detection limit (Methods S3). We estimated an infection risk curve, that is, a function relating pre-infection antibody levels to the probability of infection using a descending logistic function. For infected individuals, we extracted model-predicted antibody titres at the estimated time of infection; for uninfected individuals, we used antibody titres measured at the midpoint of the surveillance period (when 50% of observed infections had occurred within each wave. Individual exposure probability was modelled as a logistic function of the number of infected household members, and the infection likelihood incorporated a mixture of household and background community transmission. Hierarchical effects assessed variation in antibody kinetics and correlations of protection by infection history (Methods S3.2.5). Models were fitted separately for each biomarker and wave combination, and we also developed dual-biomarker models with multivariate logistic protection functions (Methods S3.3). Implementation of all models is described in Methods S4. Full model code is available at https://github.com/ccgh-idd/cop-transvir-sarscov2.

### INFERENCE OF MISSED INFECTIONS

Infections missed by PCR surveillance were identified using serojump, (18) a reversible-jump MCMC method that probabilistically infers individual infection status and timing from serological rises in spike and nucleocapsid proteins antibodies (Methods S3.1.1). The PCR-confirmed infections captured 34% and 42% incidence during the Delta and Omicron waves respectively, while the combined PCR and serologically-inferred infection captured higher incidence rates of 53% and 79% for the same periods. Of 348 participants in 52 households, 289 participants in 49 households with sufficient serological data were included; 43/289 (14%) received 1³ SARS-CoV-2 vaccine dose.

### MODEL COMPARISON AND CONTERFACTUAL ANALYSIS

Models were evaluated using Receiver Operating Characteristic Area Under the Curve (ROC AUC) and Leave-One-Out Cross-Validation (LOO-CV PSIS). Both metrics are transdimensionally comparable, depending only on predicted probabilities versus observed outcomes. The biomarker with the lowest composite predictive ranking score was selected as the optimal correlate (Methods S3.4). Using optimal correlates, we simulated 4-fold and 8-fold titre increases (representing vaccine or natural boosting) from pre-season antibody levels, and estimated population-level protection changes through absolute and relative risk reduction (Methods S3.5). These analyses assume uniform response across the population and do not account for potential heterogeneity in vaccine-induced antibody responses observed in real-world settings.(10)

## RESULTS

### LONGITUDINAL TRENDS IN IMMUNE BIOMARKERS

We analysed longitudinal antibody responses from 289 participants with 768 serological and mucosal samples collected between March 2021 and July 2022 (Figure 1). Serum pseudovirus neutralisation titres (pVNT) against the Delta increased from a median IC50 of 36 (95% CI: 6-194) before the Delta wave, to 145 (95% CI: 10-1,666) after the Delta wave. Similarly, pVNT against Omicron BA.1 increased from an IC50 of 28 (95% CI: 2–348) before the Omicron BA.1 wave, to 51 IC50 (95% CI: 8–2,115). Mucosal IgA responses showed similar patterns: Delta IgA titres increased from 32 AU/mL (95% CI: 3-370) to 51 (95% CI: 5–804) after the Delta wave, and Omicron BA.1 IgA from 11 AU/mL (95% CI: 2–111) to 47 (95% CI: 4–480) after the Omicron wave.

**Figure 1.**
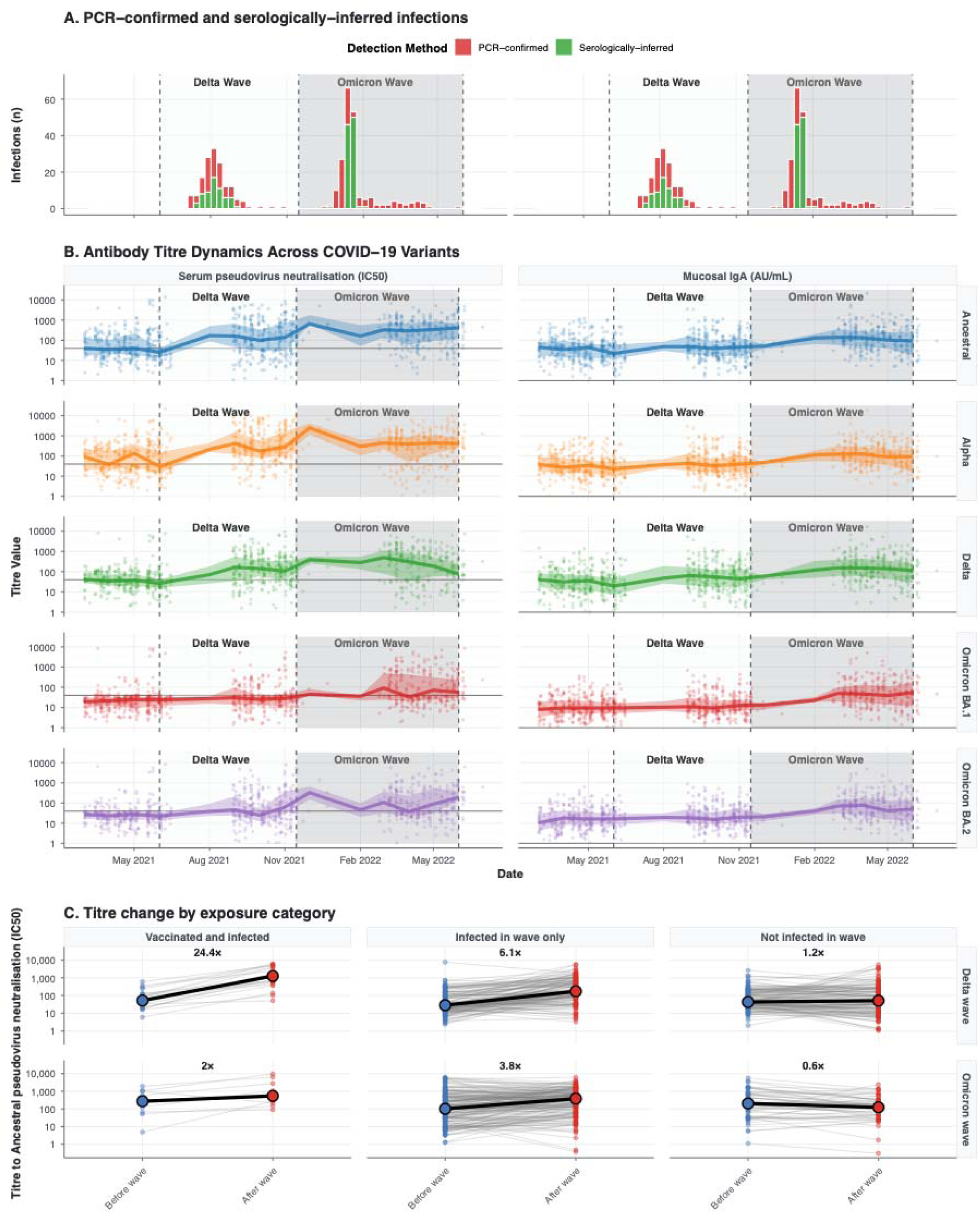
Longitudinal infection and antibody titre dynamics across SARS-CoV-2 variants during the COVID-19 pandemic in The Gambia. **A**. Infection patterns across two epidemic waves, with cases distinguished by two detection methods: PCR-confirmed (red bars) and serologically inferred (green bars). **B**. Temporal patterns of serum pseudovirus neutralisation (pVNT) and mucosal IgA antibody responses against five SARS-CoV-2 variants (Ancestral, Alpha, Delta, Omicron BA.1, and Omicron BA.2). Individual data points are point-markers, with distribution ribbons (25th-75th percentiles) and median lines illustrating population-level trends. Wave periods are indicated by background shading: Delta wave (June-November 2021) in light gray and Omicron wave (November 2021-June 2022) in darker gray, with vertical dashed lines marking wave boundaries. **C**. Titre change by exposure status for the Delta and Omicron waves (rows) for before and after the wave.

Participants with both infections and vaccination during the Delta wave experienced 24-fold rises in Ancestral pVNT after Delta infection versus 6-fold for infection alone. During the Omicron wave, this pattern reversed (2-fold versus 4-fold), suggesting an antibody ceiling effect for the infection and vaccination groups, who had higher baseline titres. Correlation analysis of baseline titres across the ten biomarkers showed similar patterns between waves (Figure S1). Mucosal IgA responses against different variants showed very high correlation (r = 0·79–0·98), indicating substantial crossreactivity. Serum pVNT showed moderate correlations (r= 0·39 – 0·80), with stronger correlations among Ancestral, Alpha and Delta variants compared to Omicron variants. Across immune compartments, correlations between serum pVNT and mucosal IgA responses, ranging from 0·26–0·50.

### RECONSTRUCTING ANTIBODY KINETICS ACROSS IMMUNE BIOMARKERS

Reconstructing antibody kinetics revealed distinct patterns of immune response and persistence across variants and biomarkers (Figure 2A). Wave-matched, variant-specific responses showed serum versus mucosal differences. During the Delta wave, infections produced substantial Delta peak pVNT-fold rises (26·8, 95% CrI 5·0–95·2) persisting 197 days >4-fold rise, while peak Delta-specific IgA responses were more modest (8·6, 95% CrI 1·7–39·3) and shorter-lived (62 days >4-fold rise). During the Omicron wave, Omicron BA.1 pVNT showed median peak fold-rises of 12·2 (95% CrI 3·3–48·5) lasting over 250 days >4-fold rise, while BA.1 IgA responses fold-rises peaked at 6.2 (95% CrI 2·4–26·8), persisting 95 days >4-fold rise. Serum pVNT boosting was lower during the Omicron versus Delta wave, whereas mucosal IgA boosting remained relatively consistent between waves. Cross-reactive boosting patterns revealed differential responses to historical and contemporary variants. For serum pVNT during the Delta wave, infections produced similar fold-rises to all previous variants (Ancestral and Alpha; median peak fold-rises of between 22·1–28·6 and 152–202 days >4-fold rise). During the Omicron wave, serum pVNT titres against BA.1 showed much higher peak fold-rises compared to other variants (Figure 2B). Posterior predictive distributions for infection, vaccination and no-exposure groups are in Figures S2–3.

**Figure 2.**
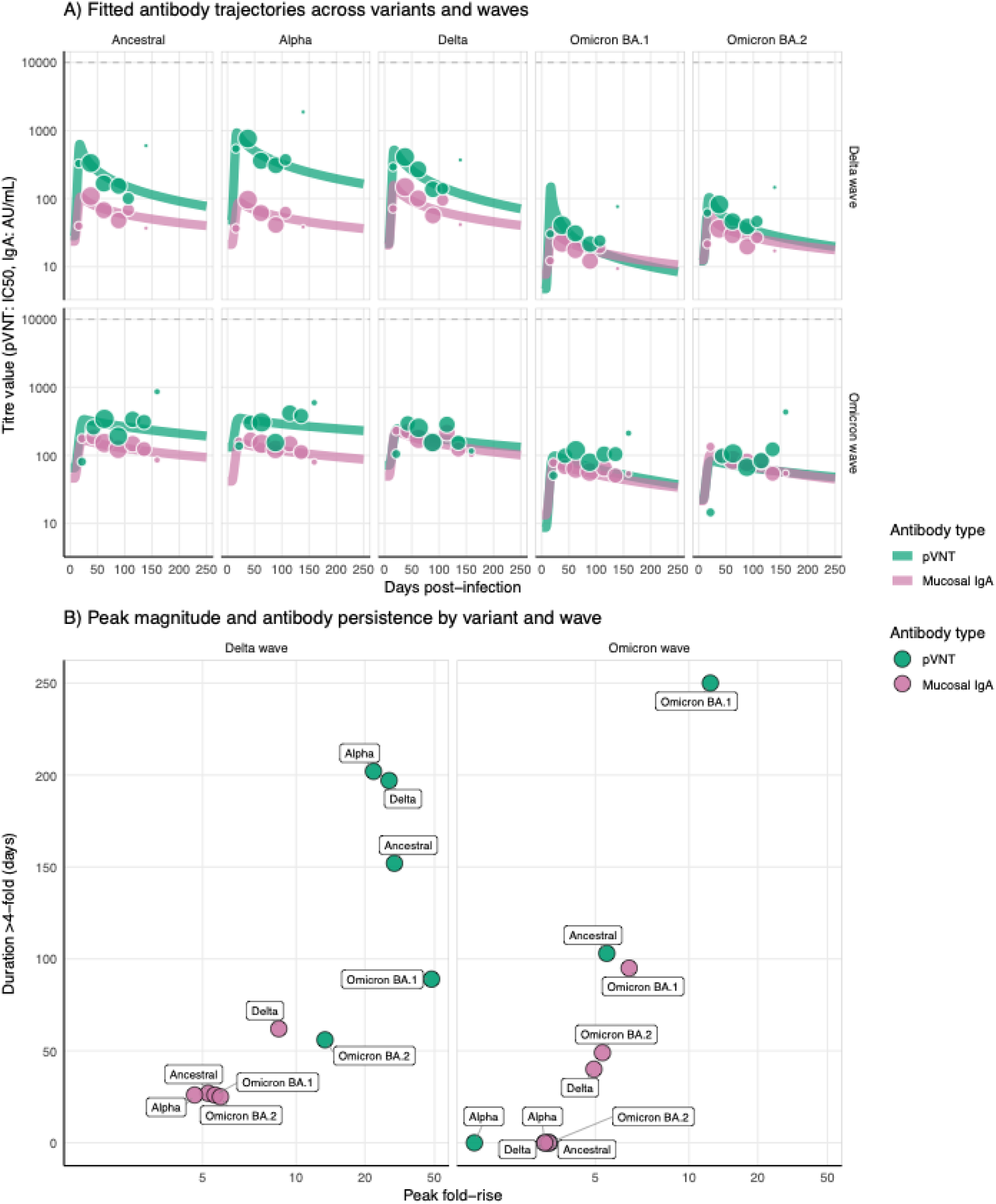
Antibody kinetics following SARS-CoV-2 infection stratified by variant and wave. (A) Fitted antibody trajectories showing titre value over time following infection for serum pseudovirus neutralization (pVNT, teal) and mucosal IgA (pink) responses across five SARS-CoV-2 variants, and two waves (Delta wave, top row; Omicron wave, bottom row). Point markers represent observed titres values from data, with size proportion to number of individuals in each bin. (B) Relationship between peak antibody magnitude (maximum fold-rise from baseline) and antibody persistence (number of days the fold rise stays above 4). Each point shows the posterior mean estimate for each variant-biomarker-wave combination.

### INFLUENCE OF INFECTION HISTORY ON ANTIBODY KINETICS

Prior infection history was associated with substantial changes in the magnitude of post-infection boosting and peak antibody titres (Figure 3). Individuals with prior infection history had elevated pre-infection titres compared to infection-naive individuals (median pre-infection Delta pVNT IC50 of 47 for previous infection vs. 10 for naïve individuals during the Delta wave, Figure 3A). First infections during the Delta wave generated substantial antibody responses, with median peak fold-rises of 59 (95% CrI 30–140) for Delta pVNT and 10 (95% CrI 5–57) for Delta mucosal IgA. In contrast, first infections during the Omicron produced lower boosting, with median peak fold-rises of 15 (95% CrI 5–71) for Omicron BA.1 pVNT and 17 (95% CrI 6–159) for Omicron BA.1 mucosal IgA. Second infections showed attenuated fold-rises in pVNT, but similar fold-rises for mucosal IgA across all variants. Individuals with ³2 prior infections demonstrated the highest pre-infection antibody levels and lowest fold-rises upon reinfection, demonstrating an antibody ceiling effect, where pre-existing immunity limits the magnitude of subsequent responses (Figure 3B).

**Figure 3.**
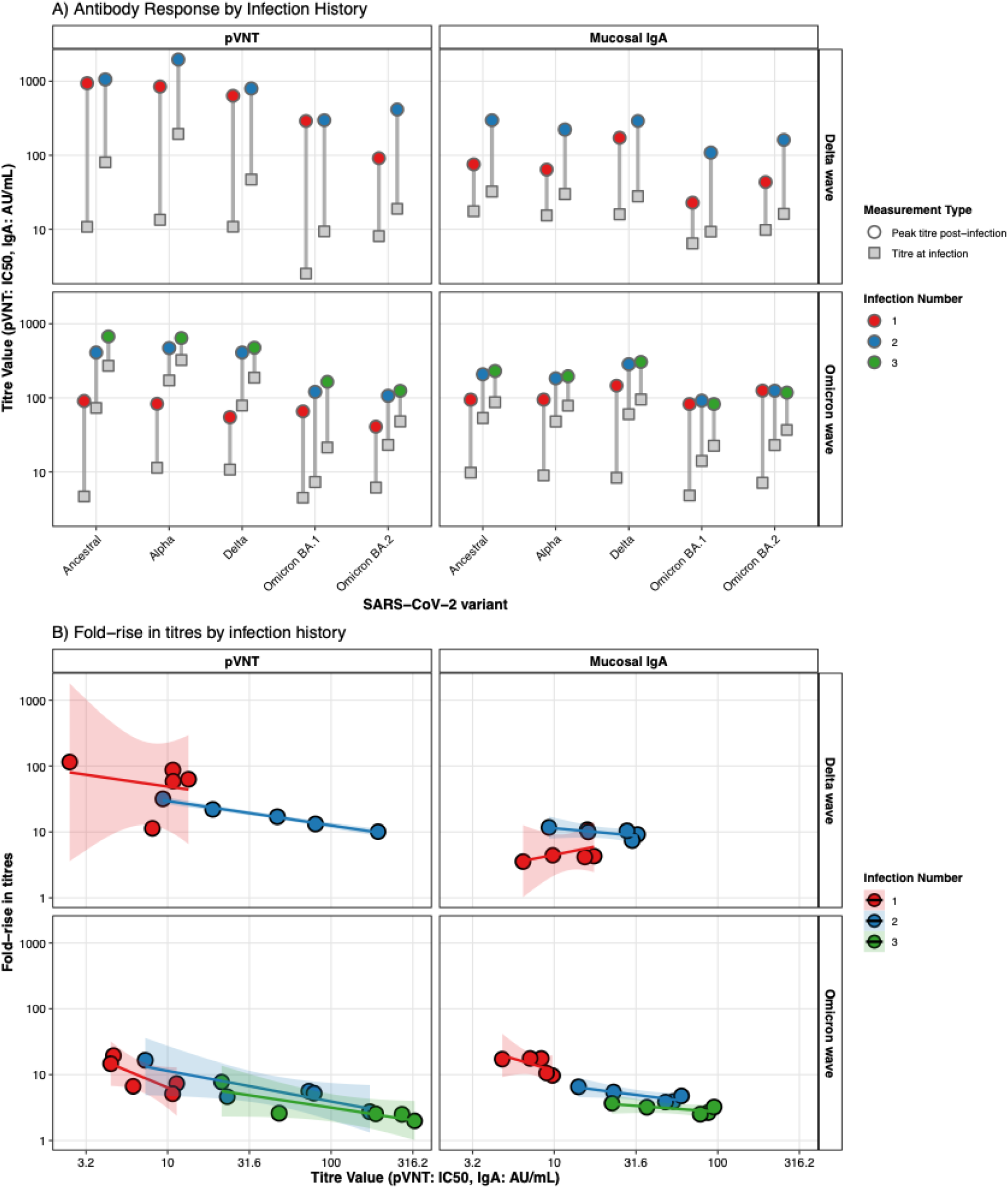
Antibody boost magnitude stratified by prior infection history. A) Gray boxes show the antibody titre at exposure across each of the five variants, two biomarkers (column panels, left, pVNT; right, mucosal IgA), across both waves (rows panels, top, Delta wave; bottom, Omicron wave). The colored circular points show the maximum titre reached post infection, split by the infection number during that wave. B) The relationship between the pre-infection titre and the fold-rise for each of the five variants, stratified by the infection history (color), wave (panel rows) and biomarker (panel columns)

To determine whether infection history independently affects boosting, we performed linear regression relating pre-infection titres to post-infection fold-rises. First infections showed highly variable boosting, whereas second and subsequent infections showed more consistent responses (Figure S4A). Baseline antibody levels explained this difference: individuals with lower baseline titres had more variable boosting than those with higher baseline titres (Figure S4B). Once we accounted for pre-infection titres, infection history no longer independently predicted boosting magnitude.

### ESTIMATED CORRELATES OF RISK AND PROTECTION

We evaluated correlates of protection against infection through a two-step approach, first comparing all individual biomarker models, then testing whether dual biomarker models (combining variant-matched serum pVNT and mucosal IgA) improved predictive accuracy. Figure 4A presents the fitted correlates of protection curves for both the Delta and Omicron waves across five variants and two biomarkers (correlates of risk curves in Figure S5; dual biomarker surfaces in Figures S6–7).

**Figure 4.**
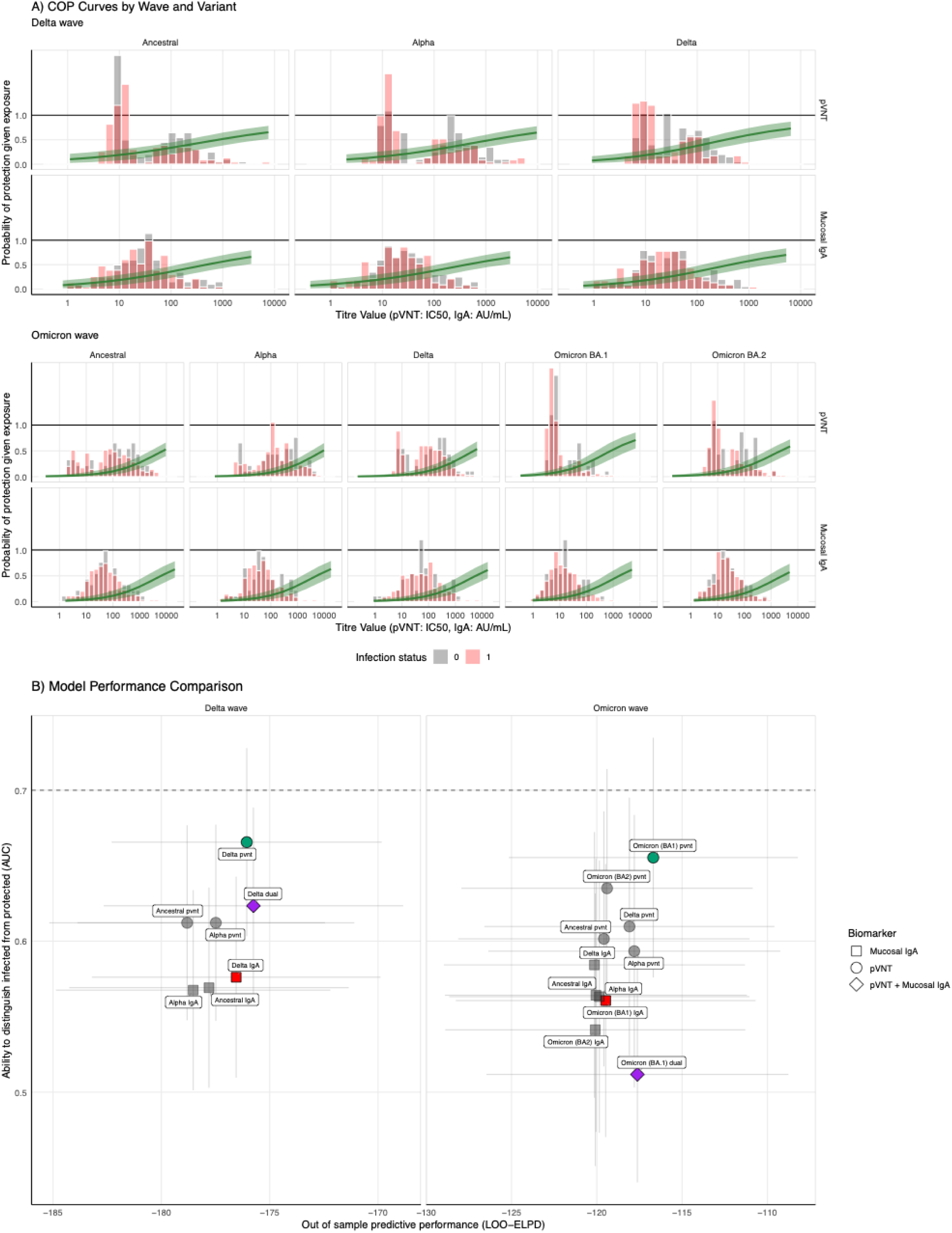
Correlation of protection analysis for SARS-CoV-2 variants across Delta and Omicron waves. (A) Fitted correlate of protection (COP) curves showing the probability of infection given exposure as a function of log_10_ antibody titre for serum pseudovirus neutralization (pVNT, top two rows) and mucosal IgA (bottom two rows) responses across five SARS-CoV-2 variants (Ancestral, Alpha, Delta, Omicron BA.1, Omicron BA.2, columns) during the Delta wave and Omicron wave. The green line is the posterior mean and the shaded region is the 95% CrI. Red bars indicate the distribution of titre values at infection for cases (infected individuals), while gray bars show the distribution of mid-season control titre values for non-cases (uninfected individuals). (B) Model performance comparison across all biomarker models for both waves. Each point represents a biomarker model, positioned according to two predictive metrics: Median P_LOO-CV ELPD on x-axis and the ROC AUC on y-axis.

Variant-matched serum neutralisation outperformed all other single biomarkers. For the Delta wave, Delta pVNT showed best discrimination (AUC = 0·66) and out-of-sample predictive accuracy, outperforming Delta mucosal IgA and all other variants. Similarly, for the Omicron wave, Omicron BA.1 pVNT demonstrated the best discrimination (AUC = 0·65) and predictive performance. For the Delta wave, although the dual biomarker model (Delta pVNT + IgA) showed comparable predictive performance to Delta pVNT alone, the single biomarker model maintained better discrimination. Therefore, we selected Delta pVNT as the optimal correlate of protection due to its greater parsimony and discriminative ability. During the Omicron wave, Omicron BA.1 pVNT sing remained optimal across all metrics. Comprehensive predictive capacity comparisons across all models are in Figure S8.

### COUNTERFACTUAL PROTECTION BY INFECTION HISTORY

Figure 5 presents model predictions comparing protection levels across alternative immunity scenarios stratified by infection history. To assess the potential impact of immunity-boosting interventions (such as vaccination of natural boosting from infection), we simulated three scenarios for each individual: observed baseline titres, and a hypothetical 4-fold and 8-fold increase from baseline. Individuals with prior infection demonstrated higher baseline probability of protection prior to each pandemic wave. (Delta wave: 19% for naive vs. 30% for 1 previous infection; Omicron wave: 14% vs. 17% vs. 24% for naïve, 1 infection and 2+ infection, respectively). Notably, despite attenuated antibody boosting in previously infected individuals, their protection gains under hypothetical boosting scenarios were comparable to naïve individuals.

**Figure 5.**
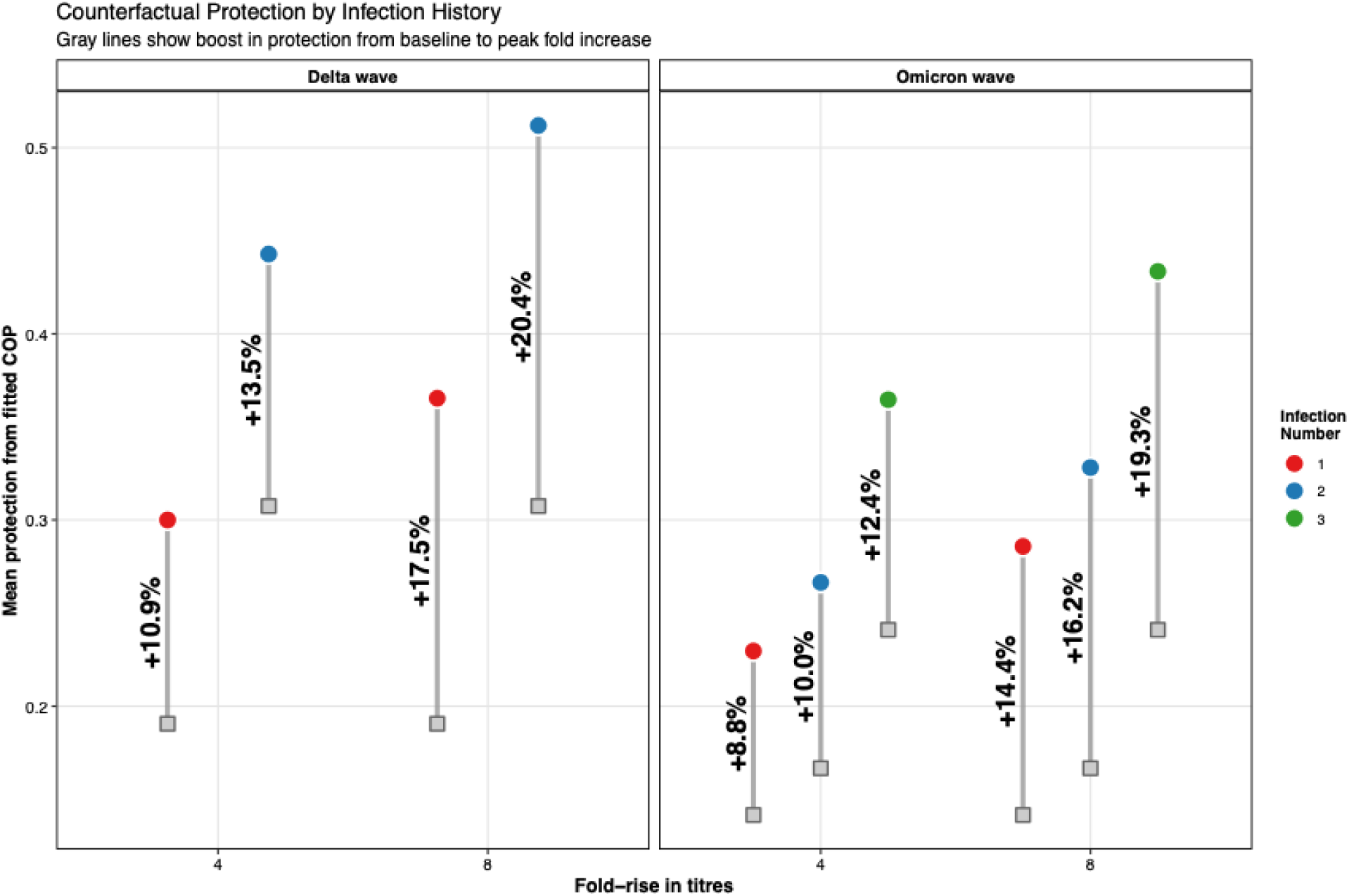
Alternative immunity scenarios stratified by infection history. Grey lines show the boost in protection from baseline (grey squares) to post-boosted titres (colored circles) for Delta (left panel) and Omicron (right panel) variants. Points represent the median protection estimate. Protection levels are stratified by infection history: naïve individuals (red), those with one prior infection (blue), and those with two or more prior infections (green).

## DISCUSSION

Our study addresses critical gaps in understanding SARS-CoV-2 immunity in African populations by characterising antibody dynamics across multiple variants in a context where natural infection predominates over vaccination. Through intensive sampling spanning the Delta and Omicron waves, we demonstrate three key findings. First, predominantly asymptomatic infections generate robust and durable antibody responses; second, that prior infection history substantially shapes both baseline immunity and subsequent boosting through immune imprinting by repeat natural infection; and third, that serum neutralising antibody titres against the circulating variant capture protection as effectively as combination of serum pVNT and mucosal IgA measurements.

Our antibody kinetics findings extend existing literature by providing multi-variant response estimates from a resource-limited setting. The median 27 peak fold-rise in Delta serum neutralising antibodies with 197 days persistence ³4-fold baseline aligns during Delta infection aligns with reports of neutralising persistence for up to 10 months with an 14·7 week half-life in predominantly mild COVID-19,(19) and a biphasic decay kinetics with a 47-day half-life in the first 6 months.(20) The high rate of asymptomatic infection may explain the more modest persistence compared to estimates of 50% protection remaining 900 days in symptomatic patients.(21) Substantially shorter mucosal IgA persistence during the Delta wave (34 days compared to 197 days for serum) agreed with findings that IgA responses were modest and diminished faster than serum IgG for the early pandemic.(22,23)

Infection history substantially influenced antibody responses, with Delta infections generating substantial back-boosting to antigenically similar variants (Ancestral, Alpha, Delta) despite high pre-infection titres (Figure 3). This aligns with observations that antibody responses typically cross-react with Ancestral spike antigens in individuals previously exposed (24,25) and reflects viral antigenic evolution and immune imprinting, where early variants cluster in antigenic space and thus share more epitopes than Omicron, which is an antigenic outlier. Attenuated responses during the Omicron wave are consistent with reports of reduced neutralising titres boosting against Omicron compared to previous variants(26,27), possibly due to Omicron variants’ preference to replicate in the upper respiratory airways, attenuating serum neutralising responses while maintaining mucosal IgA responses versus Delta.(28) Consistent with another African study, (26) predominantly asymptomatic infections generate robust, durable, and variant-responsive antibody responses with differential boosting patterns based on infection history and antigenic distance.

Critically, we found that hybrid immunity, infection followed by vaccination, produces substantially higher neutralising antibody responses than infection alone. During the Delta wave, participants with both infection and vaccination experienced 24-fold rises in Ancestral pVNT titres compared to 6-fold rises in those with infection only. This finding indicates that vaccination provides meaningful immunological benefit even in populations with extensive prior infection exposure. This has important implications for vaccination policy in high-seroprevalence African settings, suggesting vaccine deployment substantially enhances population-level immunity beyond what natural infection alone.

Single-biomarker models performed comparably to dual-biomarker models in predicting protection from protection. Variant-matched serum neutralisation provided optimal correlates of protection for both waves. Although dual biomarker models incorporating both serum neutralisation and mucosal IgA showed similar out-of-sample predictive performance, they did not substantially enhance discrimination or overall predictive accuracy, suggesting that the protective information captured by mucosal IgA is mainly contained within functional neutralising assays. While previous studies have identified neutralising antibodies as correlates of protection, (27,28) these used aggregated data across heterogeneous populations. Our household-based study with intensive surveillance enabled inference of protection curves within a well-characterised population experiencing defined exposure events. This is significant for resource-constrained settings where assay availability, cost, and complexity are critical considerations. We found that individuals mounting smaller fold-rises in antibody titres can achieve similar protection, explained by the non-linear relationship between antibody titre and protection. Individuals with higher baseline titres occupy steeper regions of the dose-response curve, where incremental increases in antibody titre translate to larger gains in protection. Thus, previously infected individual, despite smaller absolute fold-rises, derive greater protective benefit per unit titres increases versus varies individual son the curve’s flatter portion.

Our study has several notable strengths. First, the intensive longitudinal sampling enabled high-resolution characterisation of antibody kinetics rarely achieved in resource-limited settings. Further, the inclusion of multiple antibody biomarkers and the application of Bayesian statistical methods provide robust estimates of immune dynamics and protection. Second, the household-based design with defined exposure events provides more temporally precise exposure-antibody measurements than standard cohort studies. Third, minimum vaccine uptake in our cohort (14%) provides an assessment of natural infection-induced immunity without the confounding effects of heterogeneous vaccination status. Finally, the community-based sampling frame and minimal loss to follow-up ensure our findings are representative of the broader Gambian population during this period. However, this study also has several limitations. First, our study was conducted in a single region of The Gambia, potentially limiting its generalisability to other settings. Second, while we characterised antibody responses to five variants, the rapid and ongoing emergence of new antigenically drifted Omicron sublineages means these findings cannot directly predict responses to currently circulating strains, though the principles of variant-matched surveillance and antigenic distance effects remain applicable. Finally, our sample size, while substantial for a resource-limited setting, limited our power to detect small effect sizes and perform detailed subgroup analyses by age, sex, or comorbidities.

In conclusion, predominantly asymptomatic SARS-CoV-2 infections in a resource-limited African setting generate robust, durable antibody responses. Hybrid immunity from infection followed by vaccination produces substantially higher neutralising antibody levels than infection alone, supporting vaccination strategies even in highly infection-experienced populations. Further, reduced cross-protection against Omicron highlights the need for continued surveillance and variant-adapted vaccines. Finally, single-variant-matched serum neutralisation assays capture protection as effectively as complex multi-biomarker approaches, providing pragmatic, cost-effective guidance for serological surveillance in resource-constrained settings. These findings from underrepresented populations inform evidence-based pandemic preparedness strategies and underscore the essential role of diverse global immunological research in health equity.

## Data Availability

All data produced are available online at https://github.com/ccgh-idd/cop-transvir-sarscov2

## Author Contributions (CRediT)

DH: Conceptualization, Data curation, Formal analysis, Investigation, Methodology, Software, Visualization, Writing – original draft, Writing – review & editing

RW: Investigation, Writing – original draft, Writing – review & editing

NB: Investigation, Resources, Writing – review & editing

MG: Investigation, Resources, Writing – review & editing

YJJ: Investigation, Resources, Writing – review & editing

DJ: Investigation, Resources, Writing – review & editing

SJ: Investigation, Resources, Writing – review & editing

MD: Investigation, Resources, Writing – review & editing

NT: Investigation, Resources, Methodology, Writing – review & editing

BK: Conceptualization, Funding acquisition, Project administration, Supervision, Writing – review & editing

SF: Conceptualization, Funding acquisition, Supervision, Writing – review & editing

AK: Conceptualization, Supervision, Writing – original draft, Writing – review & editing

TdS: Conceptualization, Funding acquisition, Supervision, Writing – original draft, Writing – review & editing

## SUPPLEMENTARY FIGURES

**Figure S1.**
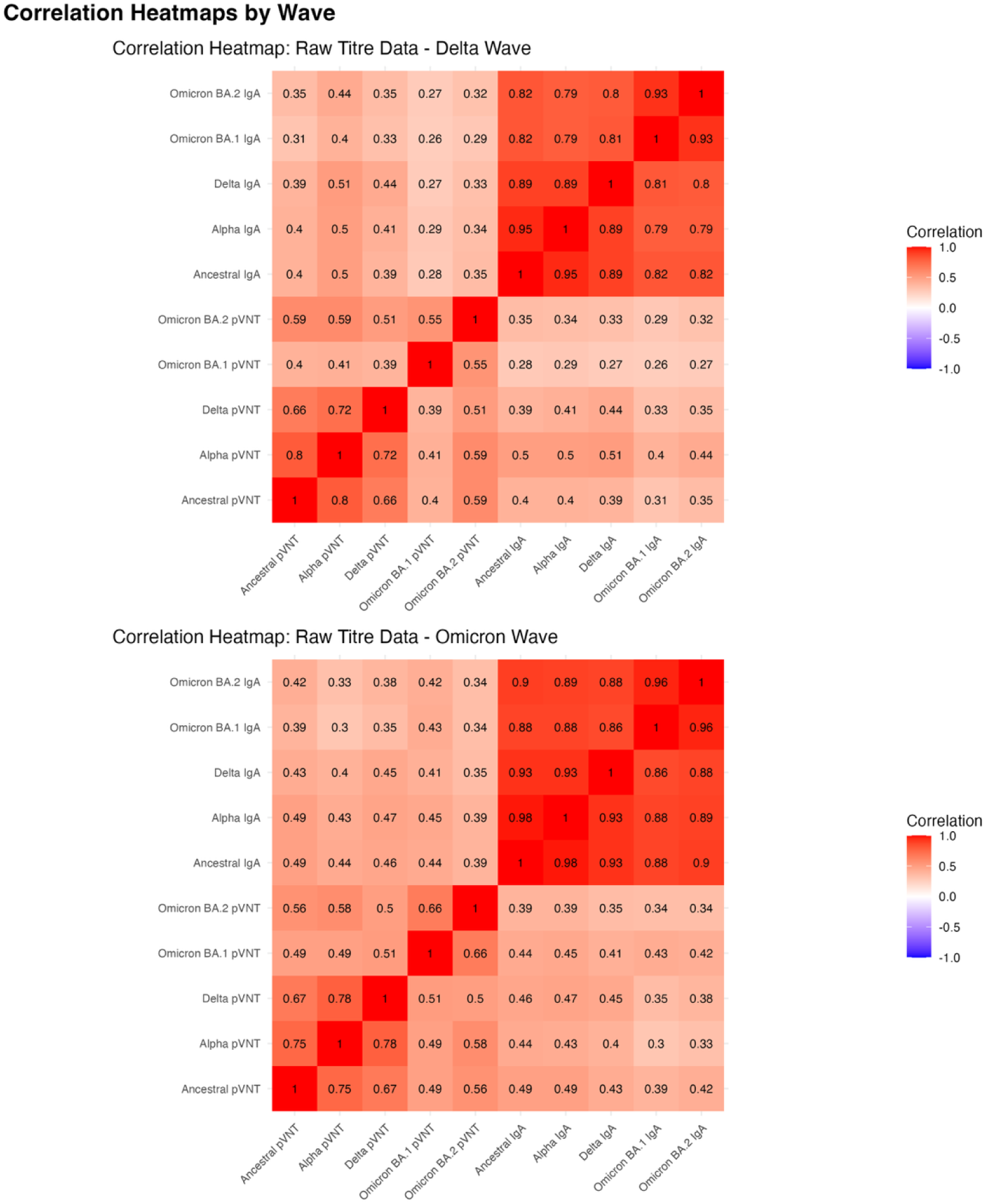
Pearsons Correlation coefficients between each of the biomarkers in the study for each study participant. Top is the correlations from the raw data across both wave, Bottom is the correlation between the inferred values from the antibody kinetic model at the point of exposure.

**Figure S2.**
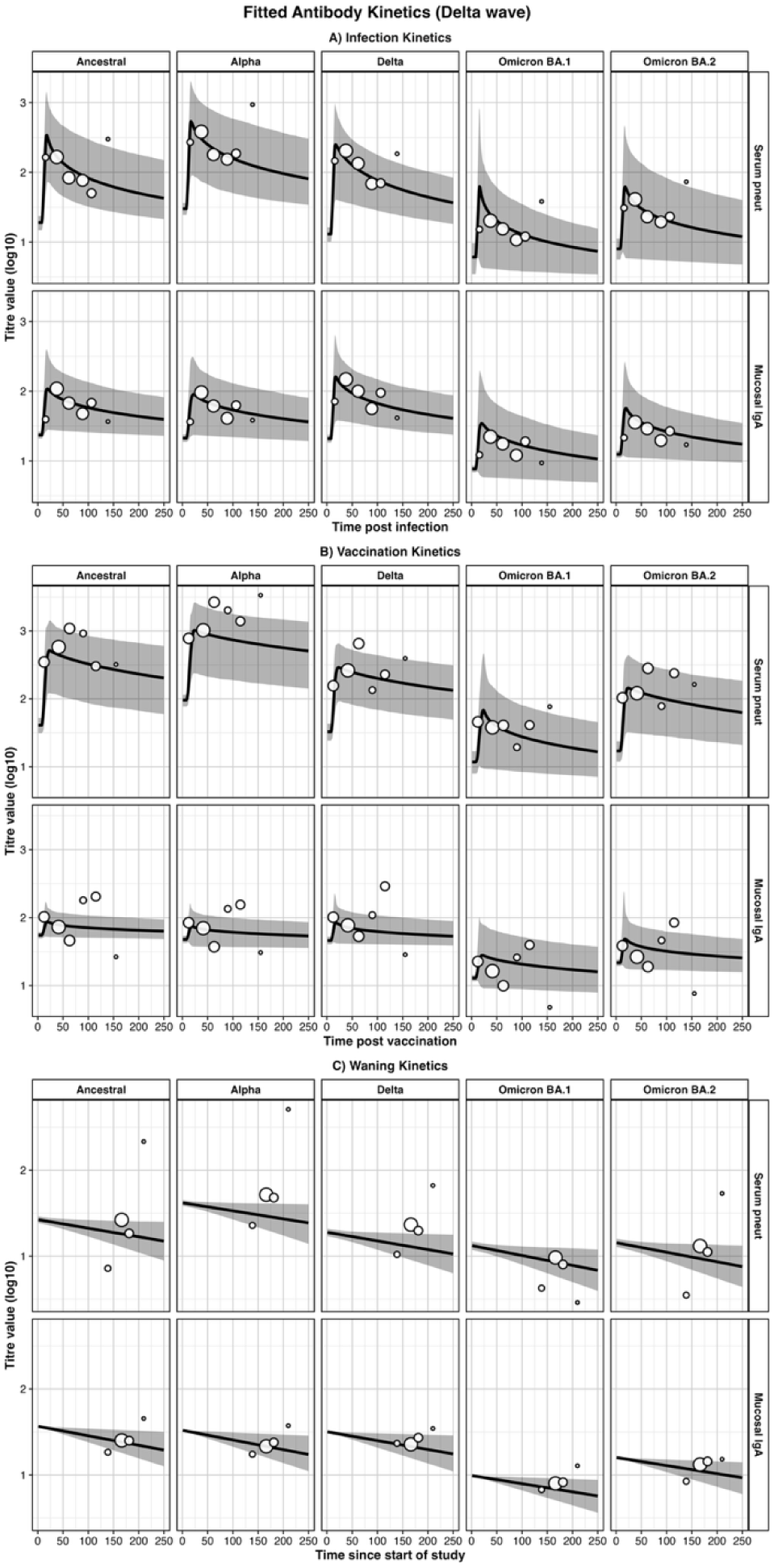
Model-fitted antibody kinetics trajectories stratified by variant, biomarker, and exposure regime (infection, vaccination, none) for the Delta wave.

**Figure S3.**
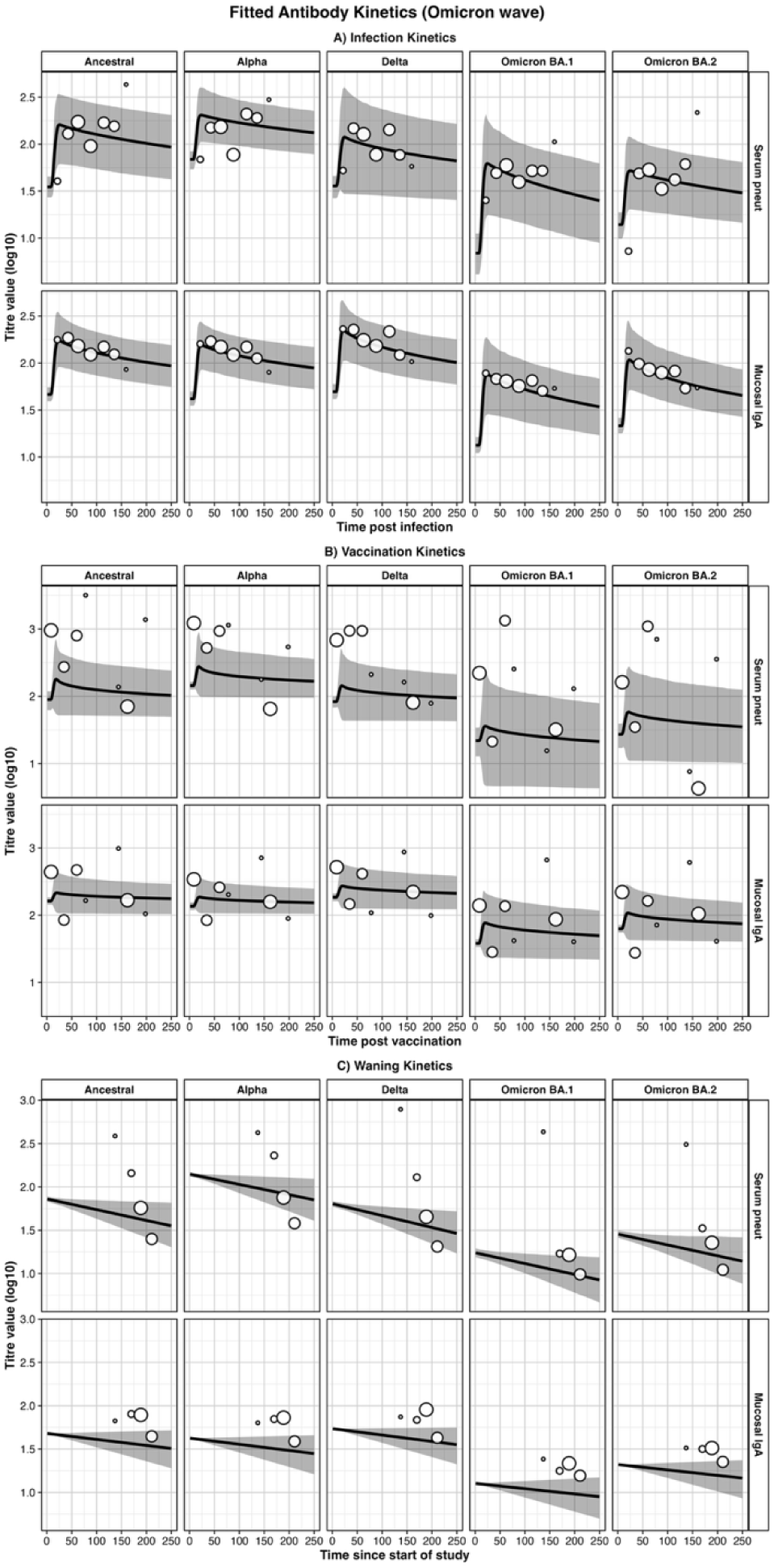
Model-fitted antibody kinetics trajectories stratified by variant, biomarker, and exposure regime (infection, vaccination, none) for the Omicron wave.

**Figure S4.**
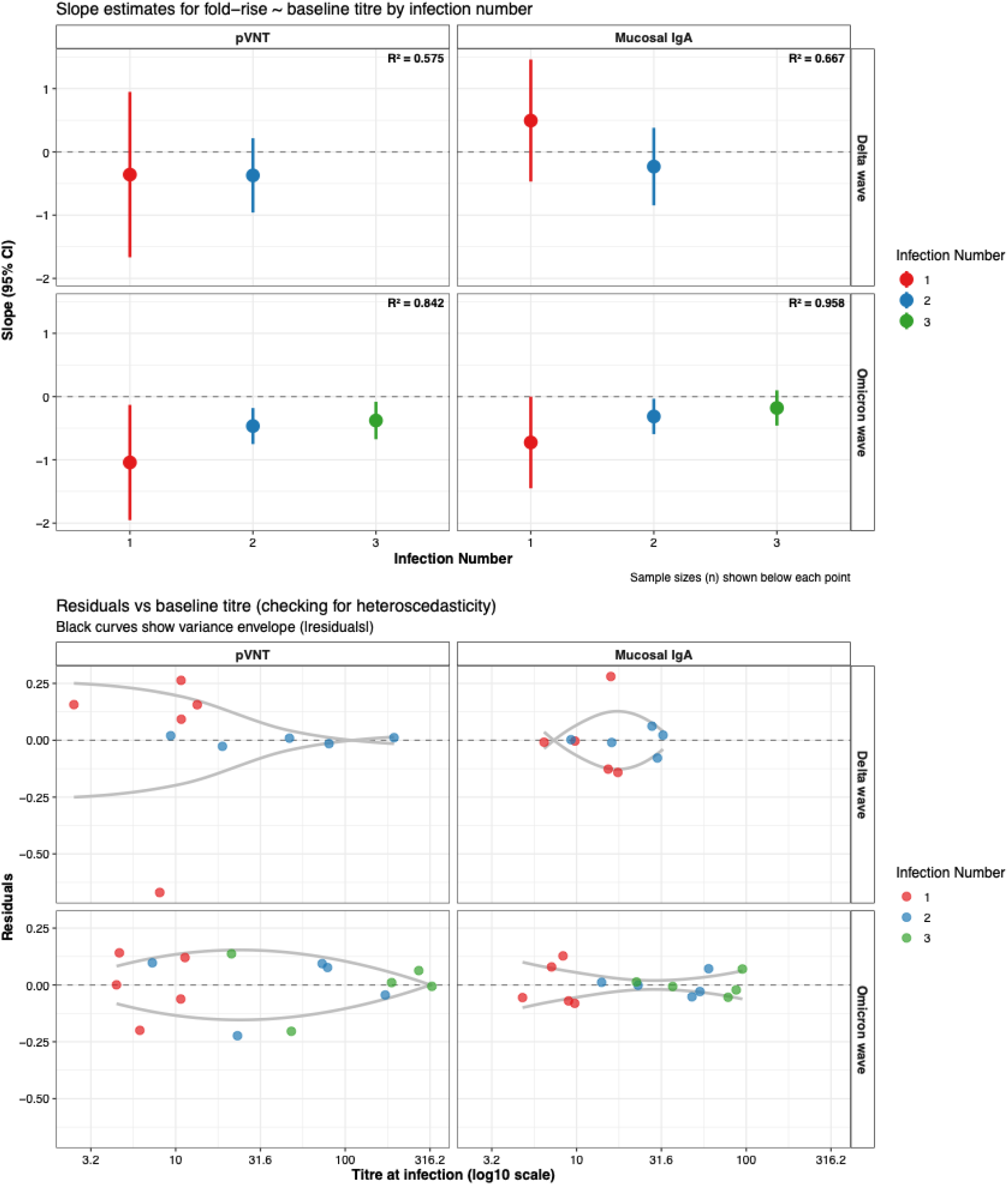
Relationship between baseline antibody titre, infection number, and immune boosting responses. (**A**) Slope estimates with 95% confidence intervals for the relationship between baseline titre and fold-rise in antibody levels following infection, stratified by infection number (first, second, or third infection). (**B**) Residual plots examining heteroscedasticity by plotting residuals from the linear regression models against baseline titre.

**Figures S5.**
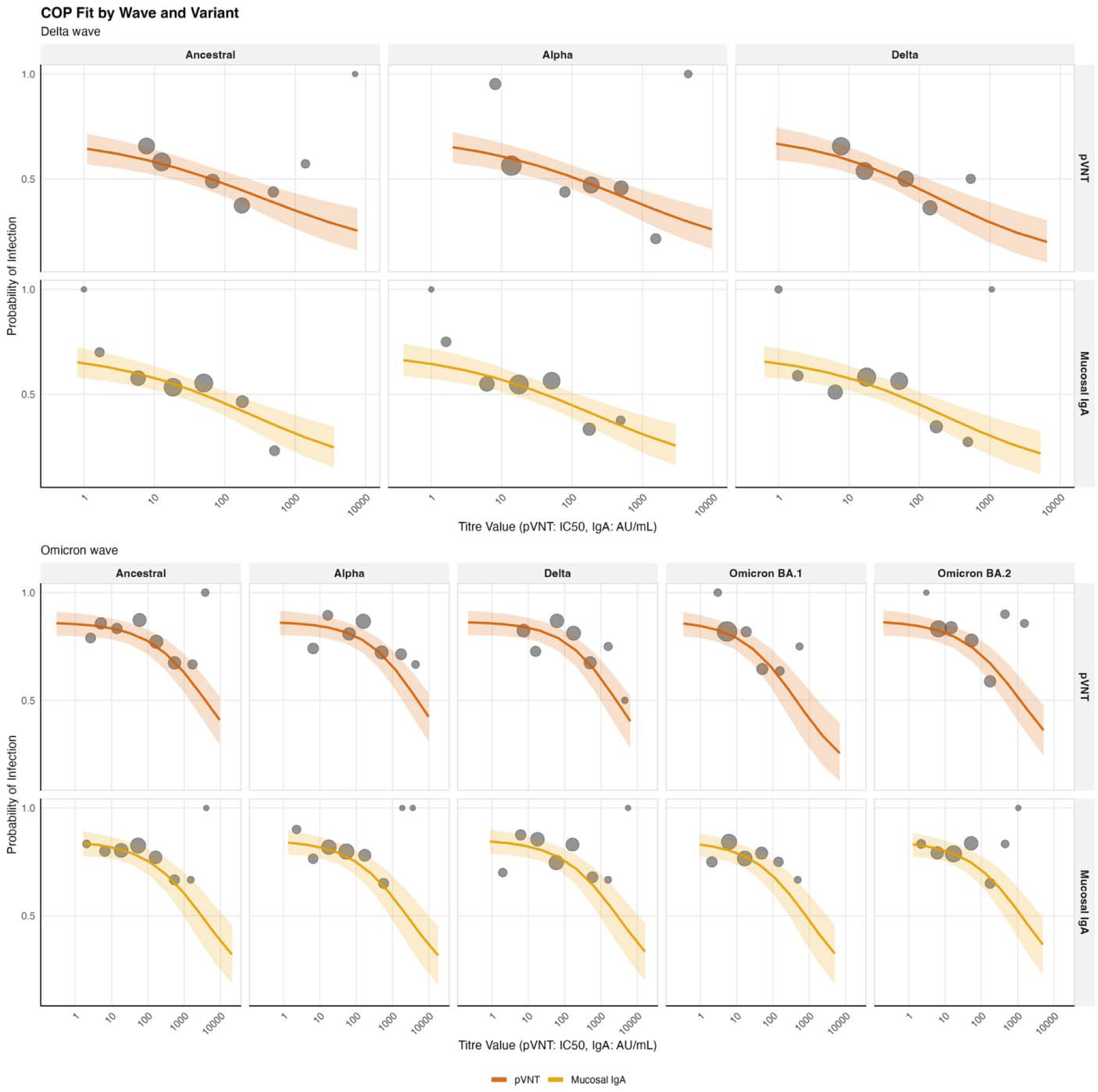
Correlates of risk curves by wave and SARS-CoV-2 variant compared to observed titre value at infection.

**Figure S6.**
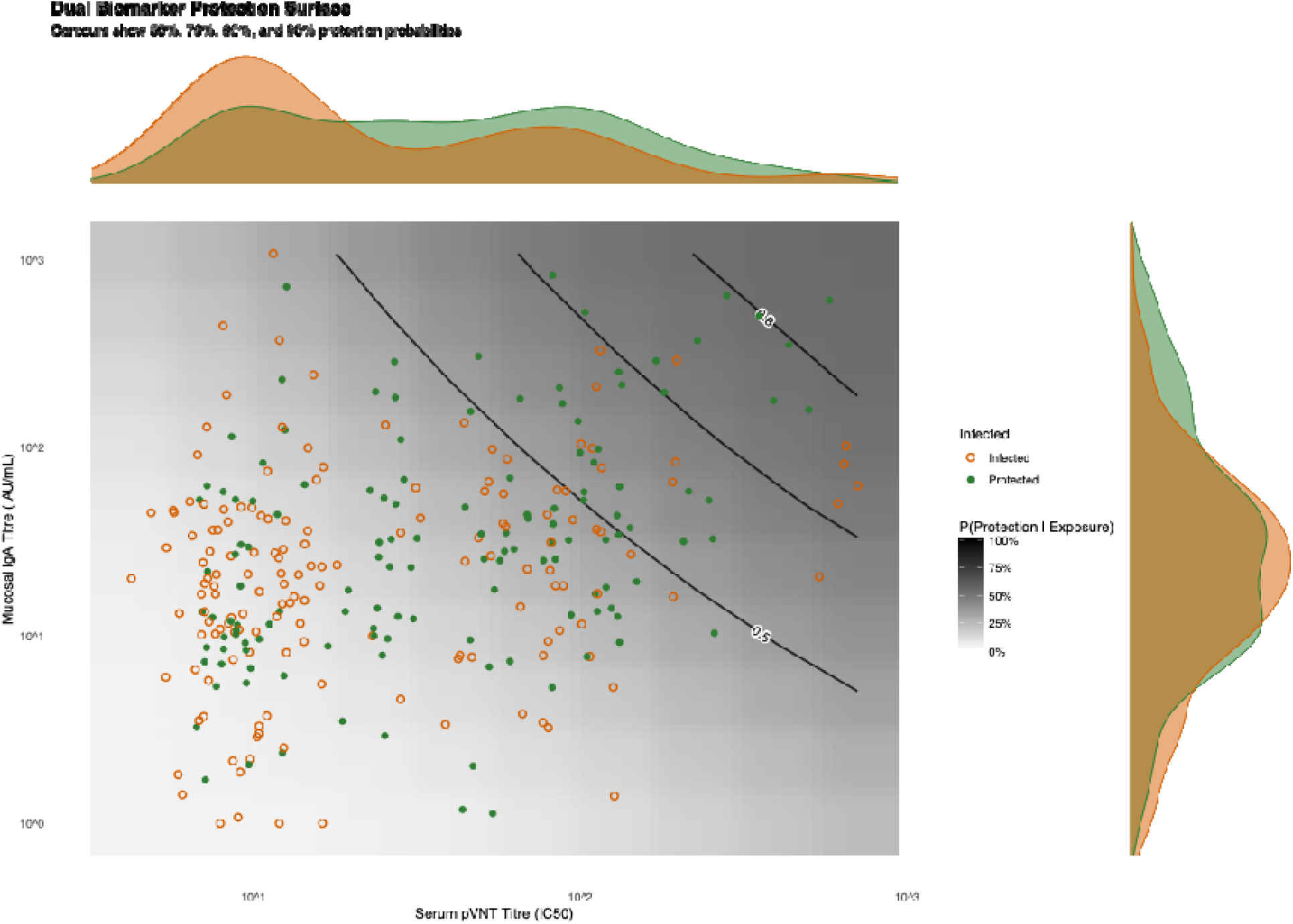
Dual biomarker Delta variant protection surface for SARS-CoV-2 infection during the Delta wave.

**Figure S7.**
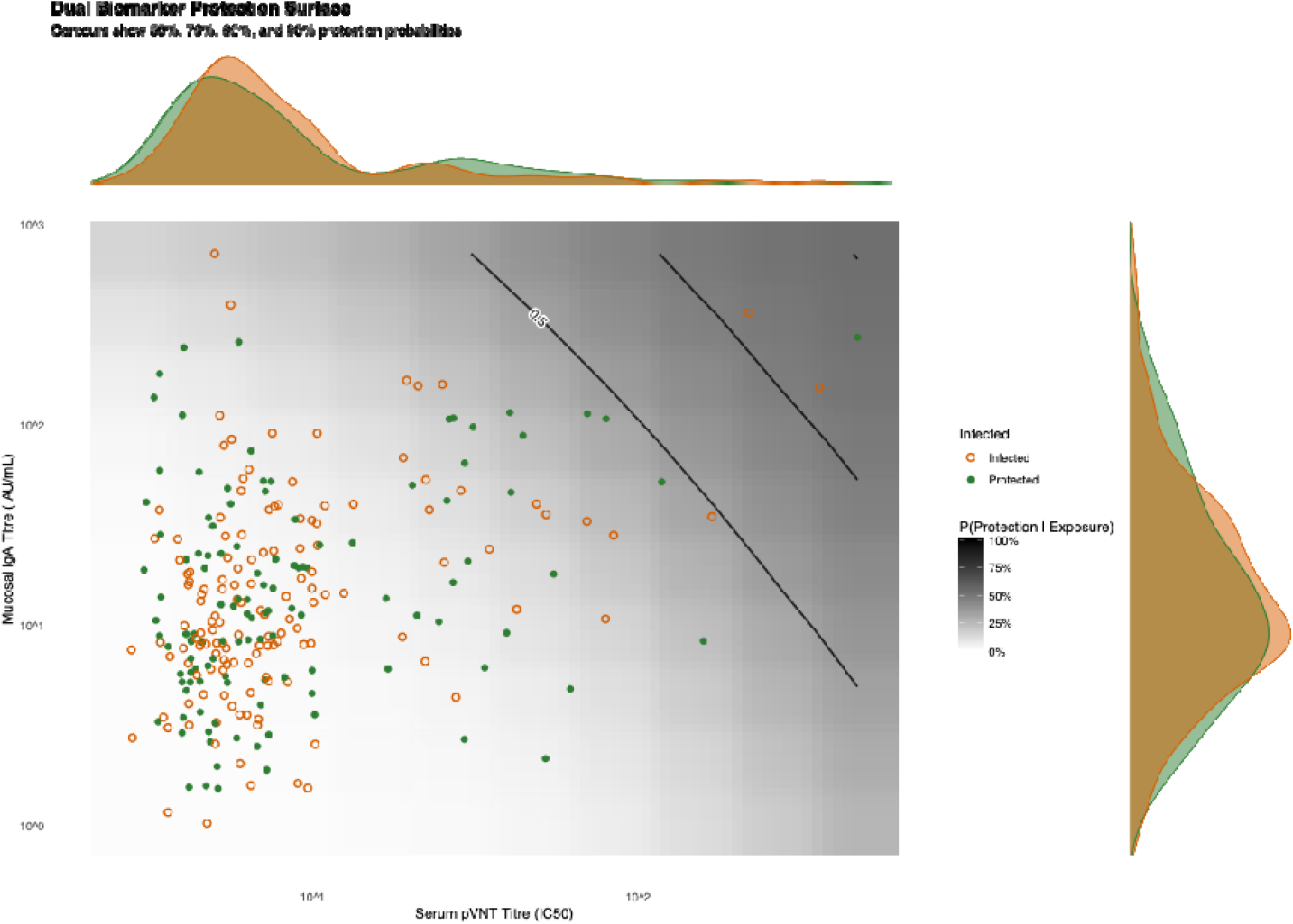
Dual biomarker Omicron BA.1 protection surface for SARS-CoV-2 infection during the Omicron wave.

**Figure S8.**
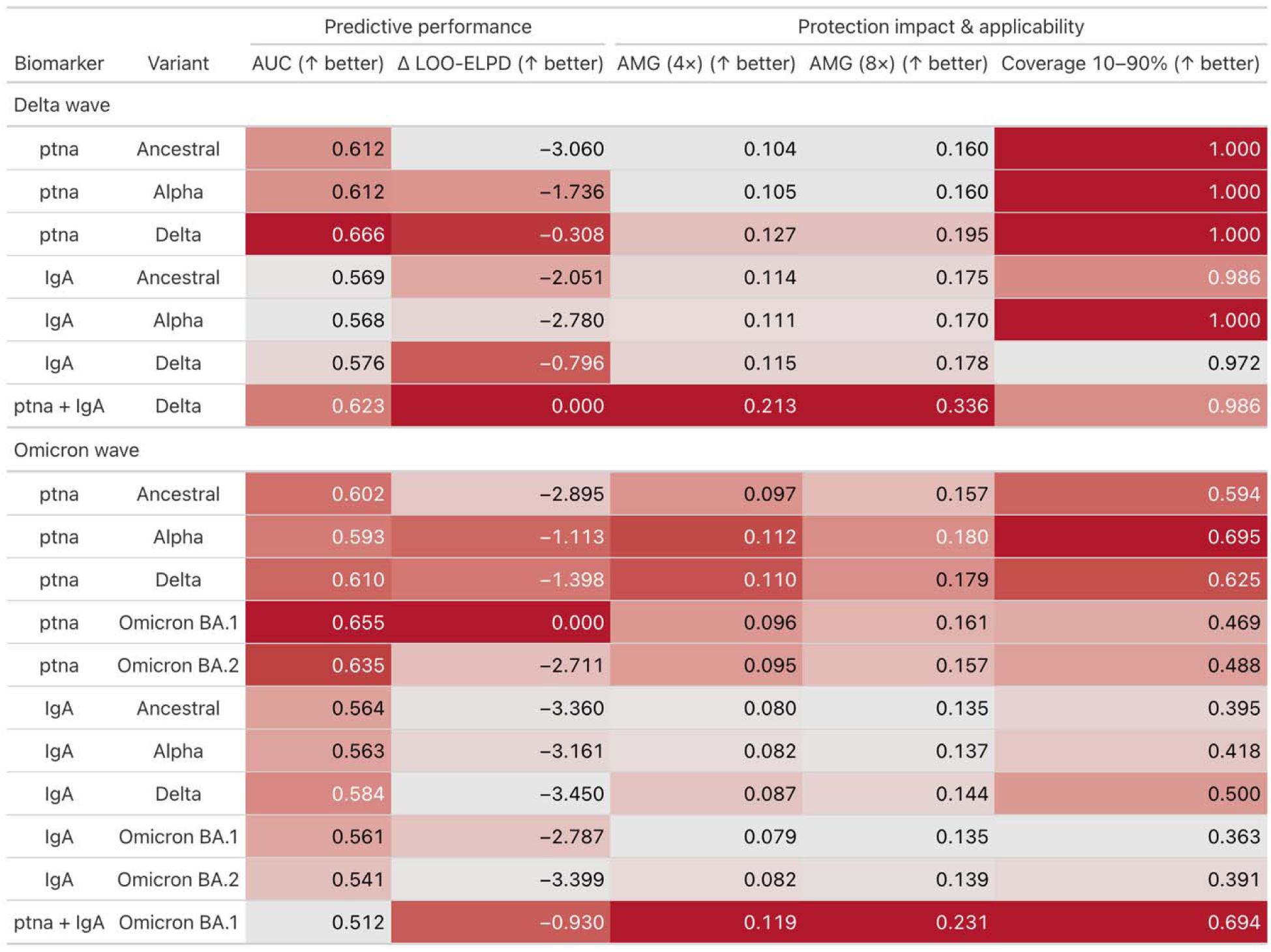
Predictive performance of single and dual biomarker models for protection against SARS-CoV-2 infection.

