## Supplementary Methods for "Impact of SARS-CoV-2 exposure history on antibody kinetics and correlates of protection in The Gambia"

Supplementary Methods to the article “Impact of SARS-CoV-2 exposure history on antibody kinetics and correlates of risk in The Gambia”

Overview

This supplementary methods document describes a Bayesian analytical framework for characterising SARS-CoV-2 antibody kinetics and identifying correlates of protection using longitudinal serological data from a cohort study in The Gambia spanning the Delta and Omicron pandemic waves. The analysis addresses three primary objectives: First, we apply the serojump reversible-jump MCMC framework to infer missed infections from serological rises in anti-spike and anti-nucleocapsid IgG antibodies, complementing PCR surveillance data. We then develop Bayesian hierarchical models to characterise the kinetics of ten distinct immunological biomarkers, capturing antibody waning, functional boosting following infection or vaccination, and measurement uncertainty. Second, we systematically identify optimal single and dual biomarker correlates of protection by extending our framework to jointly model antibody dynamics and infection risk as a function of antibody levels at exposure. Individual exposure probability is modelled through household transmission intensity, allowing us to disentangle protection from heterogeneous exposure risk. Model comparison employs two complementary metrics, ROC AUC for discriminative ability and LOO-CV PSIS for out-of-sample predictive accuracy, with biomarkers ranked by mean performance across both metrics. Third, we conduct counterfactual intervention analyses to assess the public health impact of identified correlates by simulating 4-fold and 8-fold increases in population-level antibody titres and quantifying resulting changes in protection probabilities. The models are implemented using Hamiltonian Monte Carlo via Stan. This integrated approach provides a statistical framework for understanding immune dynamics and translating serological measurements into actionable public health insights for pandemic preparedness.

TABLE OF CONTENTS

1. OUTLINE OF THE DATA

2. GOALS OF THE ANALYSIS

3. DESCRIPTION OF THE MODEL

3.1 Overview of Mathematical Structure

3.1.1. Inferring missed infections in the PCR-negative cohort from serologic rises with serojump

3.2 Bayesian Model to Infer Infection Kinetics and CoP for a Single Biomarker

3.2.1. Overview

3.2.2. Setup and notation

3.2.3. Likelihood function for the antibody kinetics model

3.2.4. Likelihood function for the protection model

3.2.5. Full posterior and priors

3.2.6. Hierarchical effects

3.3 Bayesian Model to Infer Infection Kinetics and CoP for Two Biomarkers

3.4 Model Comparison and Selection of Optimal Correlates of Protection

3.5 Counterfactual Assessment of the Best Correlate of Protection

4. IMPLEMENTATION

4.1 Software

4.2 MCMC Chain Convergence for Single and Dual Biomarkers

SUPPLEMENTARY METHODS FIGURES

Figure SM1. Correlation structure between the ttires for 10 different biomarkers for Delta Wave single-biomarker models.

Figure SM2. Correlation structure between the titres for 10 different biomarkers for Omicron Wave single-biomarker models.

Figure SM3. Incidence during each of the two waves

Figure SM4. Full schematic of the Bayesian model.

Figure SM5. Convergence diagnostics for Bayesian hierarchical models fitted to Delta wave data.

Figure SM6. Convergence diagnostics for Bayesian hierarchical models fitted to Omicron wave data

Figure SM7. Convergence diagnostics for dual-biomarker Bayesian hierarchical models

1. OUTLINE OF THE DATA

Our data consists of individual-level longitudinal serological samples taken over two waves of the COVID-19 pandemic (Delta and Omicron) from a cohort study in The Gambia.

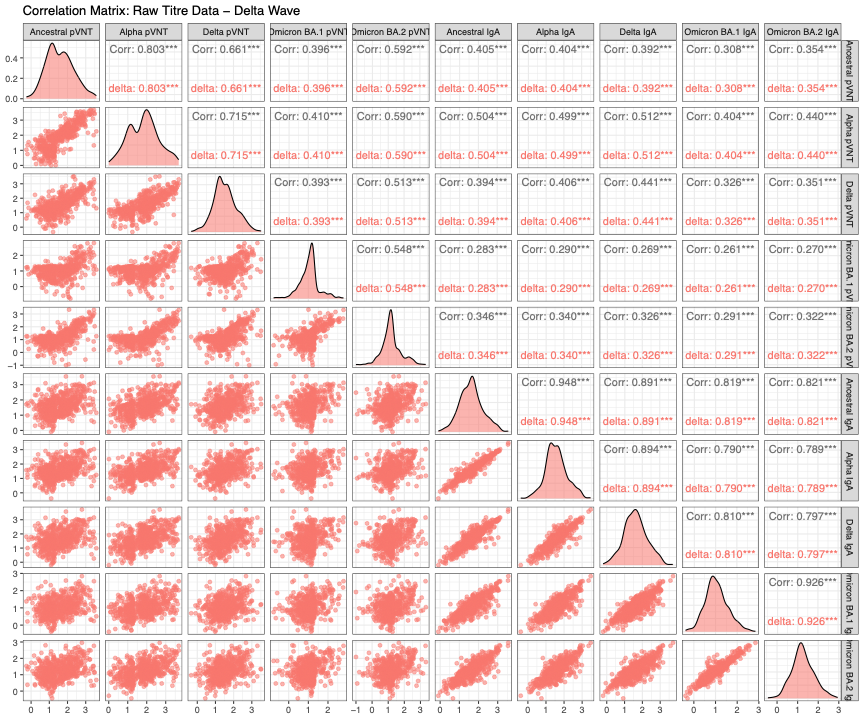

**Figure SM1. Correlation structure between the ttires for 10 different biomarkers for Delta Wave single-biomarker models. This** correlation matrix displays pairwise Pearson correlations between all biomarkers.

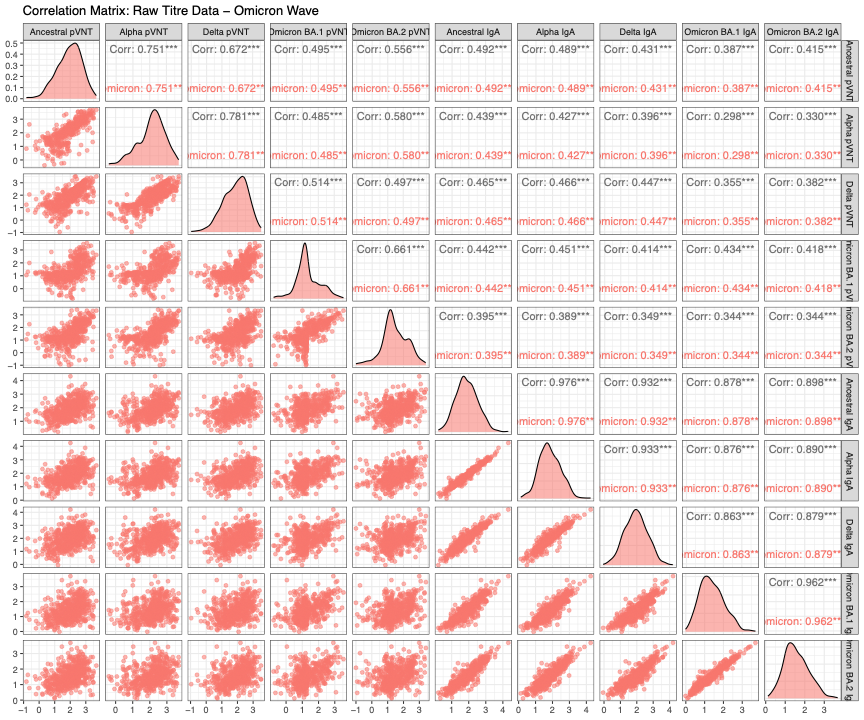

**Figure SM2. Correlation structure between the titres for 10 different biomarkers for Omicron Wave single-biomarker models**. This correlation matrix displays pairwise Pearson correlations between all biomarkers.

2. GOALS OF THE ANALYSIS

The aim of this study is to develop a comprehensive analytical framework that jointly characterises antibody kinetics and correlates of protection against SARS-CoV-2 infection using longitudinal serological data from The Gambia. By integrating individual-level immune dynamics with population-level transmission patterns, we seek to identify which immunological biomarkers best predict protection against infection. Our analysis is structured around three specific objectives:

**Objective 1: Characterize antibody kinetics and identify missed infections across two pandemic waves**

We first wish to establish a comprehensive picture of SARS-CoV-2 antibody dynamics during the Delta and Omicron waves in The Gambia and infer missed infections that PCR surveillance has missed. To achieve this objective, we first apply the serojump framework, a recently developed reversible-jump MCMC method, to infer individual infection status and timing from serological rises in anti-spike and anti-nucleocapsid IgG antibodies. This probabilistic approach accounts for uncertainty in both whether and when individuals were infected, leveraging dynamic changes in antibody trajectories rather than relying on static threshold-based heuristics. Building on these infection inferences, we then develop Bayesian hierarchical models to characterise the kinetics of ten distinct SARS-CoV-2 immunological biomarkers across both waves. These models capture three key processes: continuous antibody waning prior to exposure, functional boosting following infection or vaccination, and measurement uncertainty. By modelling antibody trajectories on the log₁₀ scale with parametric boost functions adapted from established immunological theory, we estimate population-level kinetic parameters—including waning rates, boost amplitudes, time to peak response, and decay curvatures—for each biomarker and wave combination. Critically, we fit each biomarker and wave separately, allowing us to capture how immune dynamics differ across antigenic targets and viral variants.

**Objective 2: Identify optimal single and dual biomarker correlates of protection through systematic model comparison.**

We wish to determine which immunological biomarkers, either individually or in combination, are as the most reliable correlates of protection against SARS-CoV-2 infection. A correlate of protection represents a measurable immune marker that predicts reduced infection risk given exposure. However, the optimal correlate may differ across viral variants, and combining multiple biomarkers may improve predictive accuracy by capturing distinct facets of the immune response (e.g., mucosal vs. systemic immunity, anti-spike vs. anti-nucleocapsid responses). To identify the best correlate, we extend our Bayesian hierarchical framework to jointly model antibody kinetics and infection risk. For each biomarker, we relate individual antibody levels at the time of exposure to the probability of infection using a logistic function. For cases, we use the model-predicted titre at the time of infection; for non-cases, we use a mid-season control titre (when 50% of infections have occurred) to standardise comparisons. To account for heterogeneous exposure, we model individual exposure probability as a function of household transmission intensity, allowing us to disentangle protection from exposure risk. We then systematically compare all ten single-biomarker models and selected dual-biomarker combinations using two complementary metrics: Receiver Operating Characteristic Area Under the Curve (ROC AUC), which measures discriminative ability to separate infected from protected individuals, and Leave-One-Out Cross-Validation with Pareto-Smoothed Importance Sampling (LOO-CV PSIS), which assesses out-of-sample predictive accuracy. Both metrics are transdimensionally comparable, depending only on predicted probabilities and observed outcomes,ensuring fair comparison between single- and dual-biomarker models.

**Objective 3: Assess the real-world public health impact of identified correlates through counterfactual intervention analysis**

Finally, we wish to translate our identified correlates of protection into actionable public health insights by quantifying how population-level protection would change under realistic intervention scenarios. Using the optimal single and dual biomarker correlates identified in Objective 2, we first establish baseline population-level protection by calculating the average probability of protection given exposure across all individuals based on their pre-season antibody titres. We then simulate two counterfactual scenarios representing plausible intervention strategies: a 4-fold increase in antibody titres and an 8-fold increase.

3. DESCRIPTION OF THE MODEL

3.1 OVERVIEW OF THE MATHEMATICAL STRUCTURE

We developed a Bayesian hierarchical model to describe longitudinal antibody titres in individuals with known or inferred infection and vaccination events. The model captures individual-level waning, functional boosting responses to exposures (infection or vaccination), and uses these dynamics to estimate a biomarker-based correlate of protection (CoP) via a logistic.

3.1.1. Inferring missed RSV infections in the PCR-negative cohort from serologic rises with *serojump*

To complement our primary analysis and assess the sensitivity of virological surveillance in detecting infections, we applied the *serojump* framework to the same longitudinal serological dataset from The Gambia. The *serojump* method uses reversible-jump Markov Chain Monte Carlo (RJ-MCMC) to probabilistically infer individual-level infection status and timing from changes in antibody levels over time, accounting for uncertainty in both whether and when infections occurred. Full details of the *serojump* methodology, including the antibody kinetics model specification, prior distributions, and validation studies, are provided in Hodgson et al. (2025).

Briefly, we applied serojump to infer missed infections during the Delta and Omicron waves using longitudinal IgG measurements to SARS-CoV-2 spike and nucleocapsid proteins. For each individual, *serojump* models antibody trajectories as a function of waning (prior to infection or vaccination) and boosting (following infection or vaccination) using parametric kinetic functions. The model jointly estimates population-level antibody kinetics parameters, individual infection probabilities, and the timing distribution of infections across the study period. We incorporated known vaccination dates as fixed immunological stimuli and used PCR-confirmed infection timing as an empirical prior for the epidemic curve. The analysis was run for 100,000 MCMC iterations across four chains, with convergence assessed using standard diagnostics (R̂ < 1.1, effective sample size > 1000). **Figure SM3** compares PCR-only and serojump-augmented incidence.

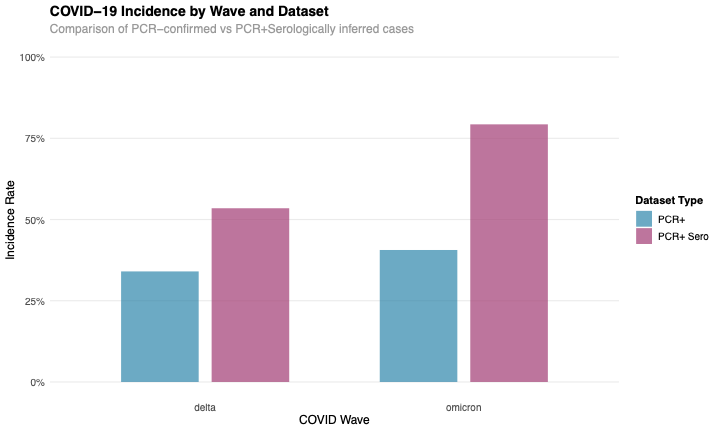

**Figure SM3. Incidence during each of the two waves,** considered using PCR+ samples only, and PCR+ samples plus serologically inferred imissed infection using *serojump*.

3.2. Bayesian model to infer infection kinetics and COP for a single biomarker

3.2.1. Overview

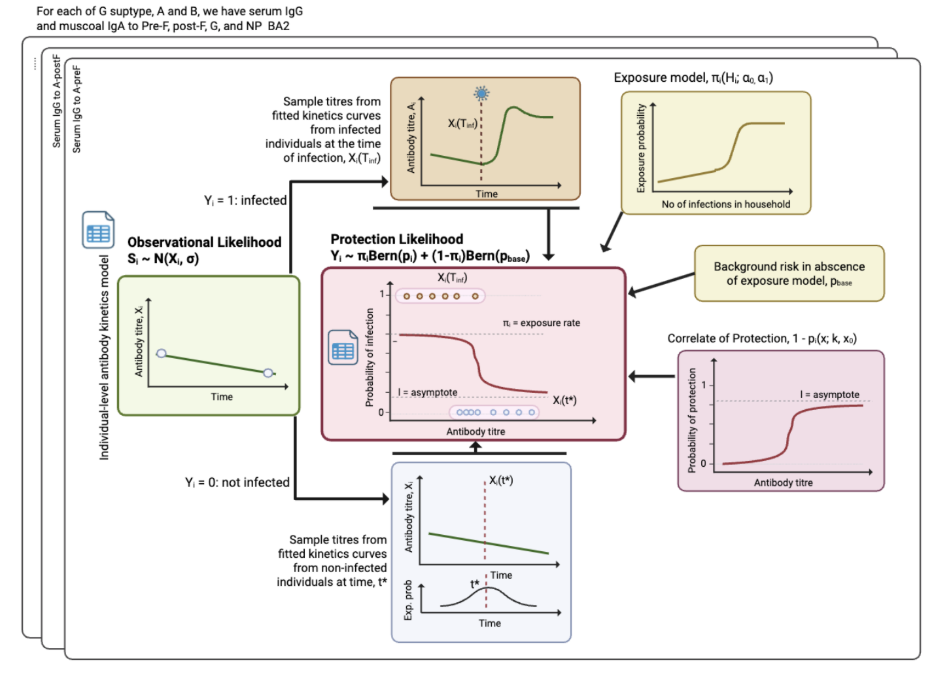

**Figure SM4. Full schematic of the Bayesian model.**

We modelled each biomarker in each wave (Delta and Omicron) by first describing how antibodies change over time for every person: before any infection, they gradually wane; if a person becomes infected, their antibodies rise in a characteristic “boost” and then slowly wane again. Using that trajectory, we predict each person’s expected titre at the study’s middle and end visits and compare these predictions to the measured values, allowing for measurement noise. To study protection, we feed a single titre into a risk model: for cases, we use the titre at their infection time; for non-cases, we use a mid-season “control” titre (the time when half of all cases have occurred). Given exposure, infection risk decreases with higher titres but never to absolute zero (we include a small residual risk). We also model each person’s chance of being exposed, which increases with the number of infected household members, and we include a small background risk to cover infections outside the measured exposure. The overall likelihood for infection combines (mixes) these exposed and background paths. Finally, we estimate everything jointly (kinetics + protection + exposure) with priors, and we allow key parameters to vary by vaccine type and host factors (age, time since vaccination) via hierarchical effects so groups can differ while still borrowing strength from one another.

3.2.2. Setup and notation

Define individuals $i \in\left\{ 1, 2, \ldots, N \right\}$, and choose a biomarker b and wave *w*, (note we fit each biomarker and wave separately, so we drop the *b*). Let log_10_ titre values at the start, middle, and end be denoted by, $S_{i, start}, S_{i, end}$, occuring at times $T_{i, start}, T_{i, end}$ and allow time to run between $T_{i, start}\leq t_{i} \leq T_{i, end}$. Finally we have an infection indicator $Y_{i} = \left\{ 0, 1 \right\}$, which indicates infection if 1, no infection otherwise, and in the case of infection, the time is given by $T_{inf}$. If there is a vaccination time, then the time is given by $T_{vac}$.

3.2.3. Likelihood function for the antibody kinetics model

3.2.3.1 Antibody kinetics functions

The log_10_ titre trajectory value, $X_{i}\left( t \right)$, at time, *t,* for each individual, *i*, is modelling through assume a structural form for the antibody kinetics, consisting of a pre-infection or continuous wane, and/or a a post-infection boost.

1. **Pre-infection wane/continuous wane:** Over the course of the study we assume that titre valeus wane linearly until the end of the study (if there is no infection), or until infection occurs. The equation the give the 10og_10_ titre trajectory value, under this process is:

$X_{i}\left( t \right) = \max\left( 0, S_{i, start} - \omega\cdot\left( t -T_{i, start} \right) \right)$,

Where the rate of waning is defined by the parameter $\omega$, and we truncate the waning at a log_10_ value of 0.

2. **Post-infection boost and post-vaccination boost:** Each infection or vaccination leads to a functional antibody increase modelled by a power-function formulation of Teunis et al. (2016). Specifically, the boost (change relative to the log_10_ titre trajectory value at infection, $X_{i}\left( T_{inf} \right)$ on the log_10_) following the time after infection is give by the equation:

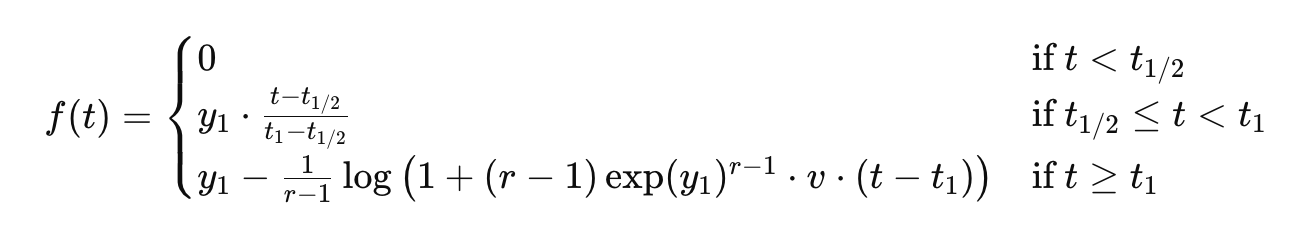
where $y_{1}$ controls the peak amplitude of the boost, $t_{1}$ is the time of the peak, *r* controls the curvature, and v is a small constant fixed to 0.001 to stabilize waning. Thus, the value of the antibody kinetics time $t_{inf}$ after infection or time $t_{vac}$ after vaccination is given by

$$X_{i}\left( t \right) = X_{i}\left( T_{inf} \right) + f\left( t_{inf} \right)$$

$$X_{i}\left( t \right) = X_{i}\left( T_{vac} \right) + f\left( t_{vac} \right)$$

From the log_10_ titre trajectory values, we can estimate the model-predicted titre values at any time *t*, for all individuals *i.* Note for individuals who have two exposures, (e.g. infection then vaccination) then the waning is calculated until the first exposure time, then the post-infection boost in calculated until the second exposure, then the post-vaccination boost is calculated until the end of the study.

3.2.3.2. Observational likelihood

Titres are modelled on the log_10_ scale with Gaussian measurement noise σ with left-censoring at Limit of Detection (LOD) (S^LOD^). The start titre contributes to the likelihood only if censored (otherwise it’s taken as given). If censored, then a latent titre value is sampled from a uniform distribution and we apply a left-censored normal CDF likelihood. For end titers likelihood is a normal distribution with mean the odel predicted titre values ($X_{i}\left( T_{i, end} \right)$) with measurement noise **σ, with left-censoring** applied i**f**$S_{i, start}$<$S^{LOD}$.

For start titres, if $S_{i, start}$ > S^LOD^, i.e. above the LOD, then there is no likelihood term (treated as fixed). Otherwise, we assume the loglikelihood for being below the LOD is given by

$$\Pr\left( S_{i, start}<S^{LOD} | S_{i}^{obs}, \sigma\right)= \Phi\left( \frac{S^{LOD}-S_{i}^{obs}}{\sigma} \right)$$

Where the latent titre value, $S_{i}^{obs}$, is has a prior density of $S_{i}^{obs} \sim U(-1.1, S^{LOD})$.

For end titres, if the observed titre value is greater than the LOD, $S_{i, end}$ > S^LOD^, then we use a normal distribution for the likelihood $S_{i, end}\sim N\left( X_{i}\left( T_{i, end} \right), \sigma\right)$. Otherwise, we apply a left-censored normal CDF as the likelihood:

$$\Pr\left( S_{i, end} <S^{LOD} | X_{i}\left( T_{i, end} \right) , \sigma\right)= \Phi\left( \frac{S^{LOD}-X_{i}\left( T_{i, end} \right)}{\sigma} \right)$$

By defining a censor indicator, $c_{i}^{(s)}\in\{0, 1\}$, for the start titre values (1: below LOD) and $c_{i}^{(e)}\in\left\{ 0, 1 \right\}$ for the end titre values. The log likelihood for the observational model is given by:

$$\log L_{i}^{obs}=c_{i}^{(s)}\left[ log\Phi\left( \frac{S^{LOD}-S_{i}^{obs}}{\sigma} \right) \right]+\left( 1-c_{i}^{\left( e \right)} \right)\log N\left( S_{i, end} | X_{i}\left( T_{i, end} \right), \sigma\right)+c_{i}^{\left( e \right)}\log\Phi\left( \frac{S^{LOD}-X_{i}\left( T_{i, end} \right)}{\sigma} \right)$$

3.2.4. Likelihood function for the Protection model

3.2.4.1 Defining the Protection model

Defining t* as the calendar time when 50% of all cases have occurred. The titre values used as inputs in the exposure-conditional protection model (see below) is given by:

$$x_{i} =\left\{ \begin{aligned} X_{i}\left( T_{inf} \right), Y_{i} = 1 \\ X_{i}\left( t^{*} \right), Y_{i} = 0 \end{aligned} \right.$$

That is, and individual is infected it is the model-predicted titre value at infection, and if not infected (but exposured, $Z_{i}=1$), it is the estimated a mid-season control titre. Using these inputs, we define the equation which defines the probability of protection against infection given exposure.

$$p_{i}^{exp} =\Pr\left( Y_{i}=1|Z_{i}=1, x_{i} \right)\mathcal{= l +}\left( 1 - \mathcal{l} \right)\frac{1}{1 + \exp\left( \beta\left( x_{i}-x_{0} \right) \right)}, \beta>0$$

It is a decreasing logistic curve with a floor value, $\mathcal{l}$, to allow residual risk at high titre values: Here, $\beta$, is the gradient of the slope at the midpoint, x_0_.

3.2.4.2 Defining the Exposure model

Let $H_{i}$ be the number of infected household members for an individual, *i*, over the season and use $H_{i}$ as proxy for the intensity of exposure for each individual. The relationship between $H_{i}$ and the individual probability of infection is assumed to be logistic:

$\pi_{i} =\Pr\left( Z_{i}=1|H_{i} \right) = \frac{1}{1 + \exp\left( \alpha_{1}\left( \alpha_{0}-H_{i} \right) \right)}$,

With $\alpha_{0}$ is the exposure level at which the gradient is steepest at $\alpha_{1}$> 0.

3.2.4.3 Mixture likelihood

Reminding ourselves of the conditional infection probability above (X), we also define:

$p_{base =}\Pr\left( Y_{i}=1|Z_{i}=0 \right)$,

Which is defined to include the possibility of infections occurring outside of the exposure model defined in this framework (e.g. infections from outside influence). It provides numerical stability during the inference process and is keep close to 0 (see priors).

With these defined, we can marganlise out the latent exposure state, $\pi_{i}$ in the likelihood through the equations:

$\Pr\left( Y_{i}=1|\left( x_{i}, H_{i} \right) = \pi_{i} p_{i}^{exp} + \left( 1-\pi_{i} \right)p_{base} \right)$,

$$\Pr\left( Y_{i}=0|\left( x_{i}, H_{i} \right)= \pi_{i}\left( 1- p_{i}^{exp} \right) + \left( 1-\pi_{i} \right)\left( 1-p_{base} \right) \right)$$

Thus the likelihood contribution is given by the equation

$$L_{i}^{mixture}\left( \theta|D \right) = \log\left( \pi_{i}\cdot Bern\left( Y_{i}|p_{i}^{exp} \right) +\left( 1-\pi_{i} \right)\cdot Bern\left( Y_{i}|p_{base} \right) \right)$$

3.2.5 Full posterior and priors

The full posterior distribution is a combination of the log likelihood below:

$$\log L\left( \theta|D \right)=\sum_{i=1}^{N} \left[ \log L_{i}^{obs}+\log L_{i}^{mixture} \right]$$

Across all individuals and plus the log of the prior densities. The prior distributions for the parameters are defined below:

3.2.5.1 Antibody kinetics model; priors and support

*Antibody Kinetics*

$$\omega\sim N\left( 0.001, 0.0005 \right)\left[ -0.1, 0.1 \right]$$

$$y_{1} \sim N\left( 0, 2 \right) \left[ -6, 6 \right]$$

$$t_{1} \sim N\left( 14, 4 \right) \left[ 7, 40 \right]$$

$$r \sim U\left( 1, 5 \right) \left[ 1, 5 \right]$$

*Likelihood*

$\sigma\sim Exponential\left( 1 \right)$

3.2.5.2 Protection model; priors and support

*Correlate of Protection model*

$$\mathcal{l\sim}U\left( 0, 0.2 \right) \left[ 0, 0.2 \right]$$

$$\beta\sim N\left( -1, 1 \right) \left[ -4, 4 \right]$$

$x_{0}\sim\frac{N\left( \left[ \max\left( S \right) + \min\left( S \right) \right] \right]}{2}, 1 \left[ \min\left( S \right), \max\left( S \right) \right]$, where S is all the titre values in the study.

*Exposure model*

$$\alpha_{1}\sim N\left( 1, 0.5 \right) \left[ 0, \right]$$

$$\alpha_{0}\sim N\left( 2, 2 \right) \left[ -5, 10 \right]$$

*Likelihood*

$$p_{base} \sim Beta\left( 1, 50 \right)$$

3.2.6. HIERARCHICAL EFFECTS

We want to assess the influence of vaccine type and host factors (age and time since vaccination) on the kinetics of antibody production. To do this, we add hierarchical effects to our parameters of interest:

Antibody kinetics

Formula for hierarchical effects on infection,

$\omega= \omega_{0} + z_{\omega}\sigma_{\omega}$, $\sigma_{\omega} \sim N\left( 0, 0.005 \right) \left[ 0, \right]$

$y_{1} = y_{0} + z_{y}\sigma_{y}$, $\sigma_{y} \sim N\left( 0, 1 \right) \left[ 0, \right]$

$r = r_{0} + z_{r}\sigma_{r}$, $\sigma_{r} \sim N\left( 0, 1 \right) \left[ 0, \right]$

Protection model

$x_{0} = x_{0,0} + z_{x}\sigma_{x}$, $\sigma_{x} \sim N\left( 0, 1 \right) \left[ 0, \right]$

$\beta= \beta_{0} + z_{\beta}\sigma_{\beta}$, $\sigma_{\beta} \sim N\left( 0, 0.3 \right) \left[ 0, \right]$

For all the above equations, z ~ *N*(0, 1).

3.3. BAYESIAN MODEL TO INFER INFECTION KINETICS AND COP FOR TWO BIOMARKERS

To determining the correlate of protection using two biomarkers, we employ the similar model as described above. This dual biomarker model assumes that the antibody kinetics functions and observational model run independently for each biomarker, $x_{i}^{1}$, $x_{i}^{2}$, with no shared effects. However, when fitting the correlate of protection model, we assume the exposure model is shared between both biomarkers (i.e. probability of exposure is independent of the biomarker assessed), and then when fitting the correlate of protection and correlate of risk, we fit a multivariate logistic regression model of the form

$$p_{i}^{exp} =\Pr\left( Y_{i}=1|Z_{i}=1, (x_{i}^{1}, x_{i}^{2}) \right)\mathcal{= l +}\left( 1 - \mathcal{l} \right)\frac{1}{1 + \exp\left( \beta_{1}\left( x_{i}^{1}-x_{0}^{1} \right)+\beta_{2}\left( x_{i}^{2}-x_{0}^{2} \right)+\beta_{int}(x_{i}^{1}-x_{0}^{1})(x_{i}^{2}-x_{0}^{2}) \right)}, \beta>0$$

Where $\beta_{1}$and $\beta_{2}$is the gradient of the slope at the midpoints $x_{0}^{1}$, $x_{0}^{2}$ for biomarkers 1 and 2. $\beta_{int}$ is the interaction term between the two biomarker to capture a non-linear relationship.

3.4. MODEL COMPARISON AND SELECTION OF OPTIMAL CORRELATES OF PROTECTION

When comparing across biomarkers and biomarker combinations to identify the best correlate of protection, we emphasize that the key metric defining an effective correlate is its predictive capacity to discriminate between infected and non-infected individuals. A strong correlate of protection should accurately predict protection outcomes based on antibody levels, both within the observed dataset and when generalizing to new observations. Therefore, we evaluated two complementary metrics to assess the predictive performance of each correlate of protection model: the Receiver Operating Characteristic Area Under the Curve (ROC AUC) and Leave-One-Out Cross-Validation with Pareto-Smoothed Importance Sampling (LOO-CV PSIS). These metrics provide distinct but complementary perspectives on model performance-discriminative ability and out-of-sample predictive accuracy, respectively.

The ROC AUC quantifies the discriminative ability of the correlate of protection curve to separate infected cases from protected cases. It can be interpreted as the probability that a randomly selected protected individual has a higher predicted protection probability than a randomly selected non-protected individual. An AUC of 1.0 indicates perfect discrimination, 0.5 indicates no discriminative ability (random guessing), and values between 0.5 and 1.0 reflect varying degrees of predictive performance. The ROC AUC is particularly valuable because it is threshold-independent and provides a single summary measure of discriminative ability across all possible classification rules. Importantly, the AUC depends only on the rank ordering of predicted probabilities relative to true protection status, making it comparable across models with different numbers of parameters or biomarker dimensions.

While the ROC AUC assesses discriminative ability on the training data, it does not directly measure how well the model generalizes to new, unseen individuals. To evaluate out-of-sample predictive performance, we employed Leave-One-Out Cross-Validation with Pareto-Smoothed Importance Sampling (LOO-CV PSIS), a Bayesian approach that estimates the expected log predictive density for new observations. Importantly, higher LOO-CV values indicate better out-of-sample predictive performance and the LOO-CV score depends only on the predicted probabilities $\pi_{i}.$and observed outcomes Yᵢ, making it comparable across models regardless of dimensionality or parameterisation.

To determine the optimal correlate of protection, we ranked all biomarker and biomarker combinations separately by ROC AUC (highest to lowest) and by LOO-CV PSIS (highest to lowest). For each model, we calculated the mean rank across these two metrics:

Mean Rank = (Rank_AUC + Rank_LOO) / 2

The biomarker or biomarker combination with the lowest mean rank was selected as the best overall correlate of protection.

3.5. CONTERFACTUAL ASSESSEMENT OF THE BEST CORRELATE OF PROTECTION

After identifying the optimal single and dual biomarker combinations as correlates of protection through the ranking framework described above, we conducted counterfactual analyses to assess the real-world public health impact of these correlates. Specifically, we estimated the population-level protection afforded by baseline antibody levels and then quantified how protection would change under hypothetical intervention scenarios that boost antibody titres prior to exposure.

To establish the baseline level of protection in the cohort, we used the selected correlate of protection model (the fitted logistic model from above) to estimate the probability of protection given exposure for each individual based on their baseline antibody titres at exposure. Then, to evaluate the potential impact of interventions that boost antibody levels, we simulated two counterfactual scenarios in which all individuals' baseline titres were artificially elevated prior to the season; a 4-fold and 8-fold increases in antibody titres, which represent biologically plausible ranges for vaccine-induced or natural boosting responses. Using these elevated antibody level we then calculate the change in the population-level of protection compared to the baseline antibody titres.

4. IMPLEMENTATION

4.1 SOFTWARE

The model is coded and fitted using HMC via stan using cmdnstanr version 0.5.3 in R version 4.2.3. A script for the stan code is given at <https://github.com/ccgh-idd/cop-transvir-sarscov2>.

4.2 MCMC CHAIN CONVERGENCE FOR SINGLE AND DUAL BIOMARKERS

The model is run for 4,000 steps, with 2,000 burn-in for four chains. The number of divergent transitions is 2/4000 (<0.1%), none of the chains hit the maximum tree depth of 10. The chains converged, with all parameters seeing a Potential Scale Reduction Factor (PSRF) of < 1.1, and mixed well (see Figure SM5–7).

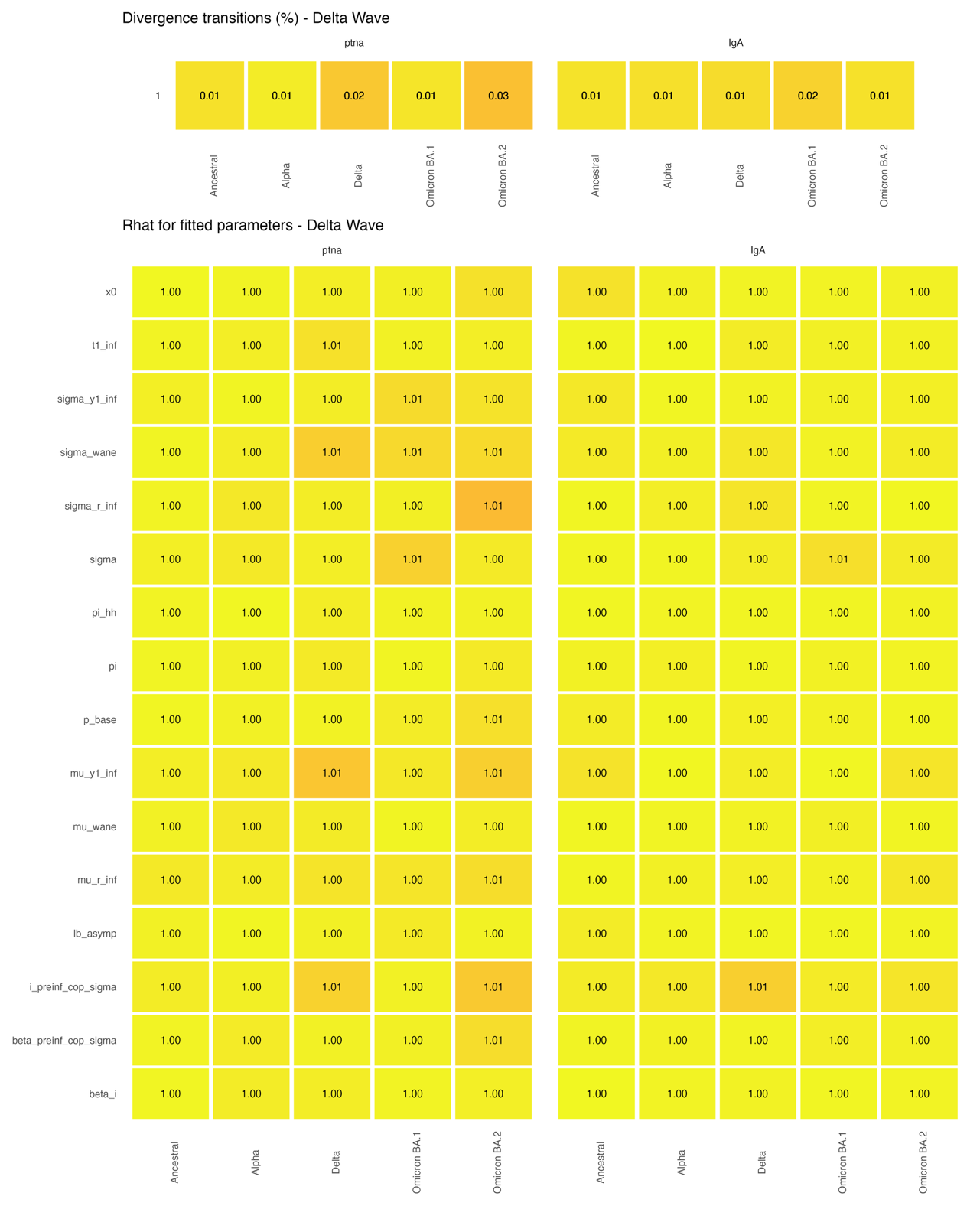

**Figure SM5. Convergence diagnostics for Bayesian hierarchical models fitted to Delta wave data**. Top panels show the proportion of divergent transitions (%) for each of ten single-biomarker models. Bottom panels display the PSRF/Gelman-Rubin statistic, for fitted parameters in each model. Parameters include: x0 (median protective titre threshold), t1_inf (time to peak antibody response post-infection), sigma_y1_inf (hierarchical standard deviation for boost amplitude), sigma_wane (hierarchical standard deviation for waning rate), sigma_r_inf (hierarchical standard deviation for response curvature), sigma (measurement noise), pi_hh (household exposure slope parameter), pi (exposure intercept parameter), p_base (baseline infection probability), mu_y1_inf (mean boost amplitude), mu_wane (mean waning rate), mu_r_inf (mean response curvature), lb_asymp (floor parameter for residual risk), i_preinf_cop_sigma (pre-infection correlate of protection hierarchical standard deviation), beta_preinf_cop_sigma (correlate of protection slope hierarchical standard deviation), and beta_i (individual-level correlate of protection slopes).

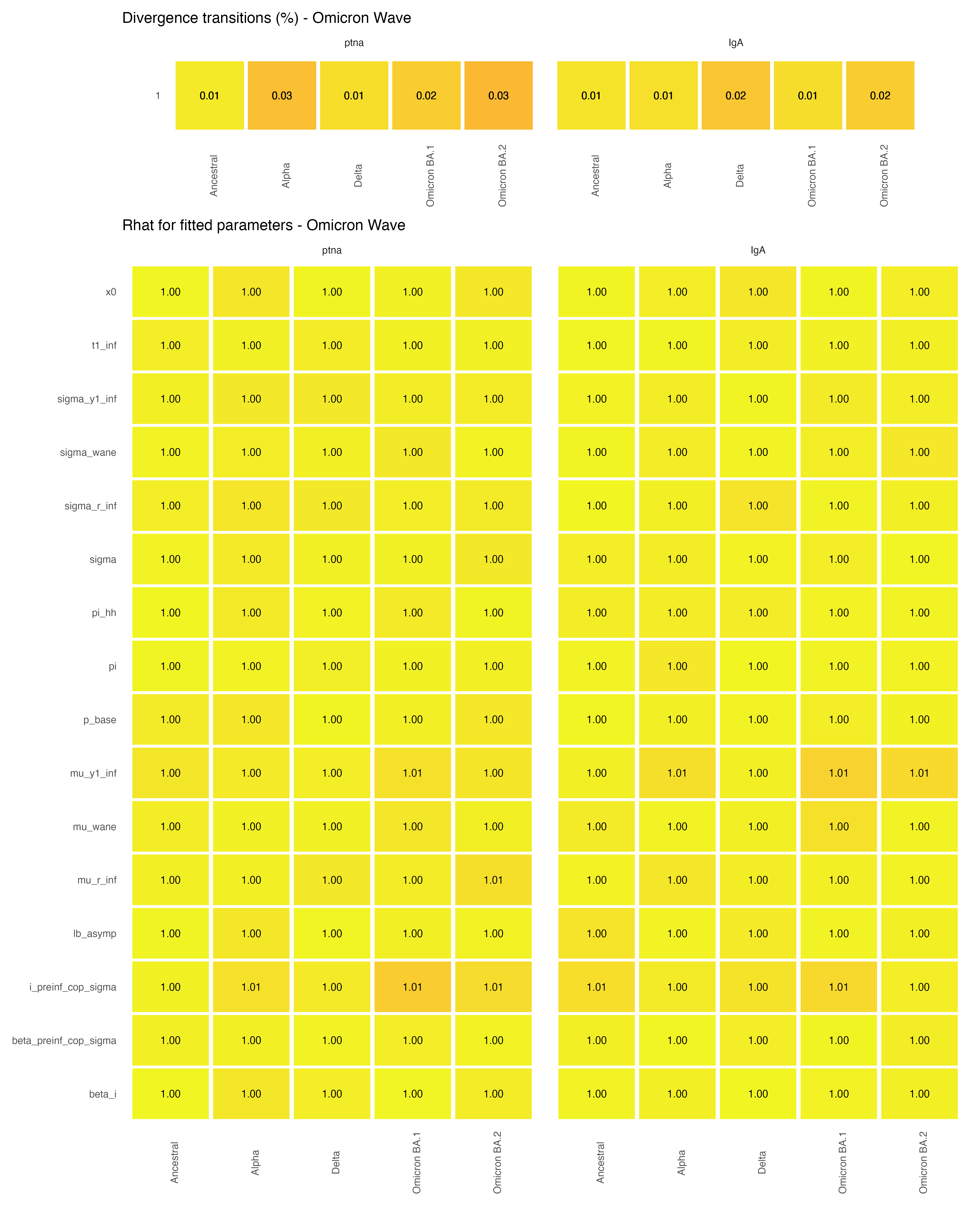

**Figure SM6. Convergence diagnostics for Bayesian hierarchical models fitted to Omicron wave data.** Top panels show the proportion of divergent transitions (%) for each of ten single-biomarker models. Bottom panels display the PSRF/Gelman-Rubin statistic, for fitted parameters in each model. Parameters include: x0 (median protective titre threshold), t1_inf (time to peak antibody response post-infection), sigma_y1_inf (hierarchical standard deviation for boost amplitude), sigma_wane (hierarchical standard deviation for waning rate), sigma_r_inf (hierarchical standard deviation for response curvature), sigma (measurement noise), pi_hh (household exposure slope parameter), pi (exposure intercept parameter), p_base (baseline infection probability), mu_y1_inf (mean boost amplitude), mu_wane (mean waning rate), mu_r_inf (mean response curvature), lb_asymp (floor parameter for residual risk), i_preinf_cop_sigma (pre-infection correlate of protection hierarchical standard deviation), beta_preinf_cop_sigma (correlate of protection slope hierarchical standard deviation), and beta_i (individual-level correlate of protection slopes).

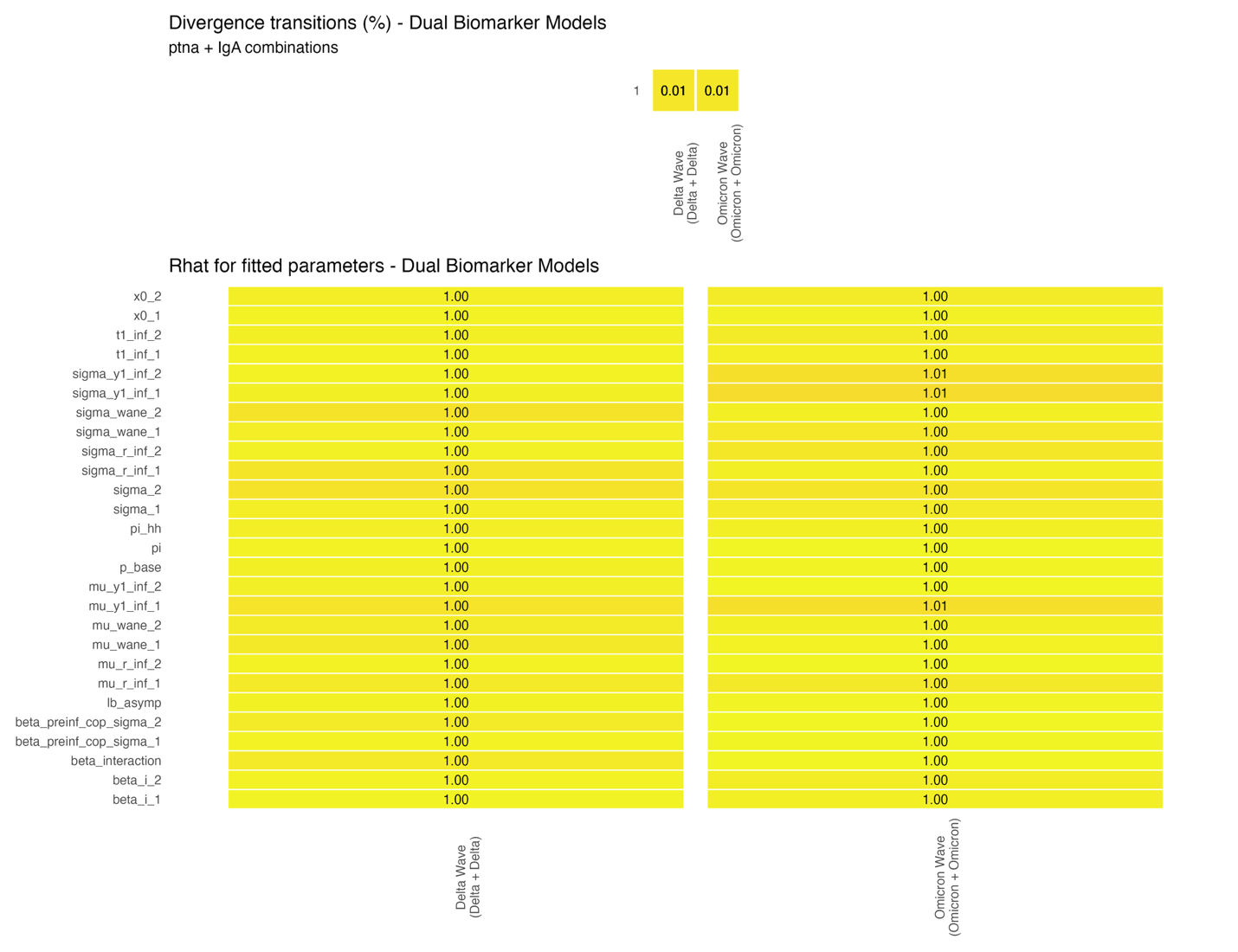

**Figure SM7. Convergence diagnostics for dual-biomarker Bayesian hierarchical models.** Top panel shows the proportion of divergent transitions (%) for two dual-biomarker models combining ptna (pre-fusion trimeric nucleoprotein antibodies) and IgA (immunoglobulin A) responses: Delta Wave model (Delta + Delta variant-specific antibodies) and Omicron Wave model (Omicron + Omicron variant-specific antibodies). Bottom panels display the PSRF/Gelman-Rubin statistic, for fitted parameters in each model.
